# Modeling optimal deployment strategies for Nipah vaccines and monoclonal antibodies

**DOI:** 10.1101/2025.10.31.25339251

**Authors:** Oscar Cortes-Azuero, Carolin Vegvari, Ellie Sutcliffe, Emma Roney, Danny Scarponi, Christinah Mukandavire, Simon Cauchemez, Emily Gurley, Henrik Salje

## Abstract

Nipah virus (NiV) is endemic in *Pteropus* bats across South and Southeast Asia. Spillover into human populations results in usually fatal disease with the potential for human-to-human spread, which has led to NiV being classified by WHO as a priority pathogen. NiV vaccines and monoclonal antibodies (mAbs) are in development. However, it remains unclear how these medical countermeasures could be used most effectively. Here, we consider three different outbreak scenarios: limited, short-lived outbreaks as seen almost annually in Bangladesh and India; livestock amplified outbreaks as seen in Malaysia in 1998; and a third theoretical scenario of an extended outbreak cause by an evolved NiV strain with increased transmissibility. For each scenario, we considered the potential impact of a vaccine when distributed through either proactive or reactive campaigns, with a hypothetical single-dose vaccine providing 90% protection against infection. We found that, in limited scenarios, the rare nature of spillovers means proactive vaccination would have a limited impact (0.21 deaths [95%CI: 0.08-0.43] averted per 10,000 doses over 30 years) and require 1.2 million doses. Reactive vaccination would also have a limited impact (0.27 deaths [95%CI: 0-2.75] averted per 10,000 doses over 30 years, 31,000 doses required, equivalent to 0.9 deaths [95%CI: 0-1] averted overall) due to the limited number of transmission generations. The use of a stockpile of 10,000 therapeutic mAb regimens that could prevent death in hospitalized patients could increase impact to 49 deaths (95%CI: 23-87) averted over 30 years. Reactive campaigns were the most effective in livestock-amplified outbreaks (2.2 deaths [95%CI: 1.6- 3] averted per 10,000 doses, 630,000 doses used in total) and extended outbreaks (467 deaths [95%CI: 293-463] averted per 10,000 doses, 690,000 doses used in total) but were highly dependent on delays to vaccine delivery. These findings suggest mAbs as the most impactful medical countermeasure under current epidemiology, with vaccines playing a significant role to limit the burden from NiV in livestock amplified and in theoretical extended outbreaks. However, the impact of both medical countermeasures will rely heavily on rapid distribution.

## Introduction

Nipah virus (NiV) is a zoonotic pathogen that circulates in *Pteropus* bat populations across South and Southeast Asia, with an estimated case fatality rate of up to 75% in humans (*1*). Pathways of transmission include consumption of contaminated raw date palm sap, contact with infected domestic animals, and human-to-human transmission (*2*). NiV was first isolated in 1999 after an outbreak among pig farm workers in Malaysia, where pigs played a key role in amplifying viral transmission (*2, 3*). Since then, all recorded outbreaks have been sporadic and localized, with Bangladesh registering outbreaks almost every year since 2001 (*4*). There are currently no approved treatments or vaccines, and NiV has been recognized as a pathogen posing a serious pandemic risk by the World Health Organization (*5*).

As of August 2025, four NiV candidates are undergoing or have recently completed Phase I clinical trials (*6*). Two viral-vectored candidates, PHV02 developed by Public Health Vaccines and ChAdOx1 NipahB developed at the University of Oxford, are due to start Phase II trials in Bangladesh (*7, 8*). Additionally, one monoclonal antibody (mAb) candidate, m102.4 completed Phase I trials in 2020 (*9*). Another candidate, MBP1F5 is due to start Phase 2 clinical trials in Bangladesh and India in 2026 (*10*). Though the development of these technologies is promising, previous studies have suggested that, primarily due to the sporadic nature and limited size of most NiV outbreaks as we know them, Phase III clinical trials would be practically unfeasible (*11*). These challenges also mean that it is unclear how best to use these medical countermeasures once approved.

Mathematical models can offer a pathway to design and evaluate the potential impact of medical countermeasure deployment strategies in different epidemiological contexts. Here we develop a framework informed by past NiV outbreaks to simulate NiV transmission, overlay context-appropriate vaccine and mAb deployment strategies, and compare their potential impact. We consider three epidemiological contexts: a limited scenario, corresponding to the most frequently observed outbreaks in Bangladesh, resulting in few cases from a spillover event; a livestock amplified scenario, corresponding to the 1998-1999 outbreak across pig farms in Malaysia; and an extended scenario, where we consider the possibility of the emergence of a more transmissible NiV strain, that would result in many more infections.

## Methods

We developed mathematical models to simulate NiV outbreaks according to three different scenarios: 1) limited outbreaks, corresponding to outbreaks triggered by spillover events from bats, and resulting in few cases, as observed in Bangladesh and India; 2) livestock amplified outbreaks, corresponding to the outbreak reported in Malaysia among pig farm workers in 1998-1999, where NiV transmission was amplified by livestock populations, resulting in hundreds of Nipah cases; and 3) extended outbreaks, in which we consider the possible emergence of a NiV strain with higher human-to-human transmissibility, resulting in widespread viral circulation. We conducted simulations over 30-year periods to estimate the long-term impact of medical countermeasures in reducing burden.

### Limited outbreaks

We extracted parameters informing NiV transmission or natural history of disease from published literature analyzing case data from NiV outbreaks in Bangladesh from 2001-2014 (Table 1), and we simulated human-to-human transmission chains with a branching process model, starting from an initial zoonotic NiV case. For each case, we drew the number of secondary cases from a negative binomial distribution with reproduction number R^L^=0.33 and dispersion parameter k=0.06 (*1, 11*). For each secondary NiV case, we drew the delay between symptom onset in the infector and transmission of the virus from a discretized gamma distribution with mean 4 days and standard deviation 2 days (*11*), and the incubation period in the secondary case from a discretized gamma distribution with mean 10 days and standard deviation 2 days (*11*). NiV cases were hospitalized with a probability of 0.9 (*1*), and the delay between disease onset and hospitalization was drawn from a discretized gamma distribution with mean 5 days and standard deviation 2 days (*11*). Similarly, hospitalizations resulted in death with a probability of 0.46/0.9 (equivalent to 0.51) in secondary cases and of 0.86 in the index case (*12*), and we drew the delay between hospitalization and death from a discretized gamma distribution with mean 2 days and standard deviation 1 day (*11*). We simulated each outbreak until the end of the transmission chain. As observed in Bangladesh, we assumed that limited outbreaks had an annual probability of occurrence of 0.8 (*4*).

**Table 1:**
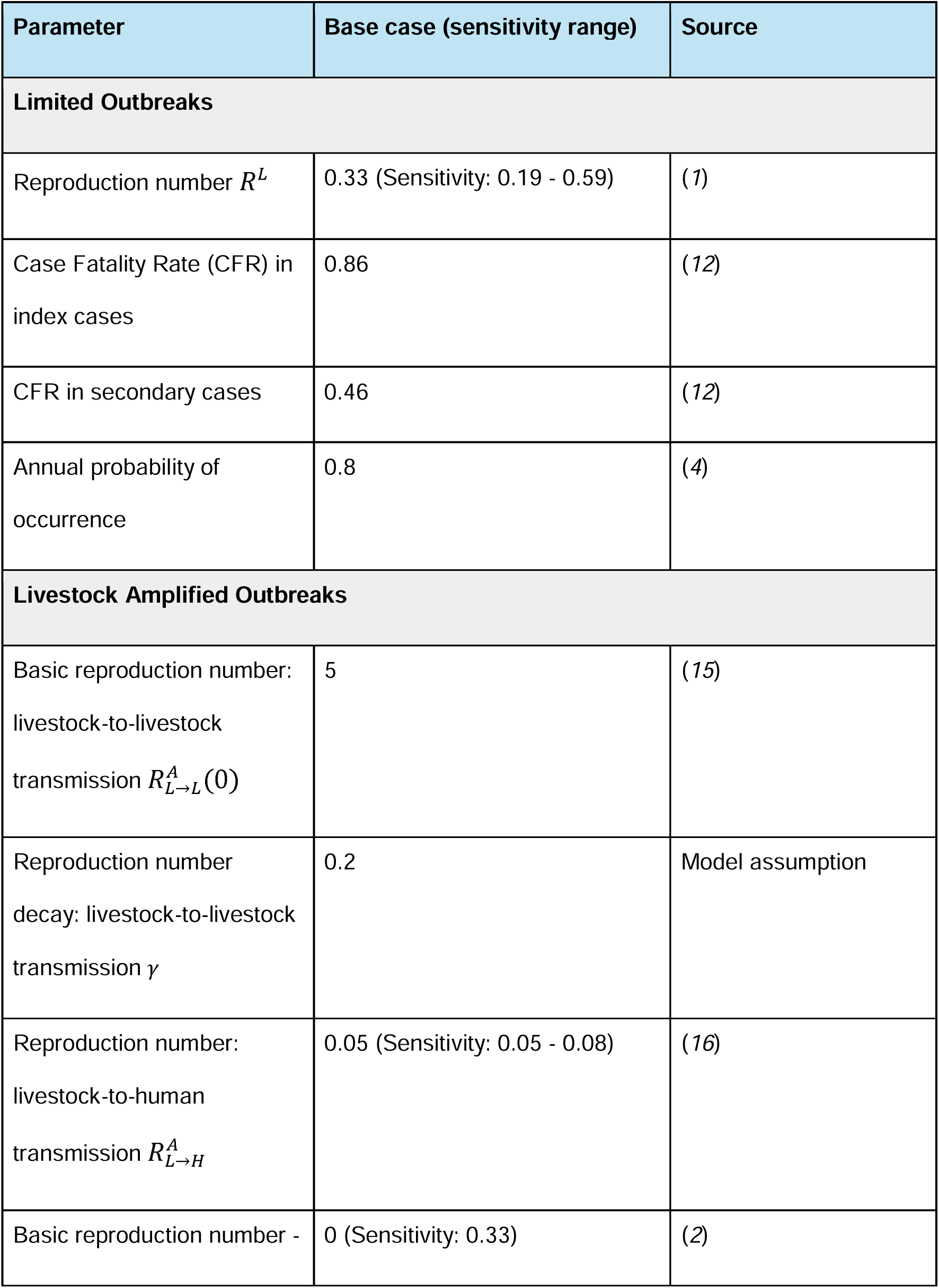
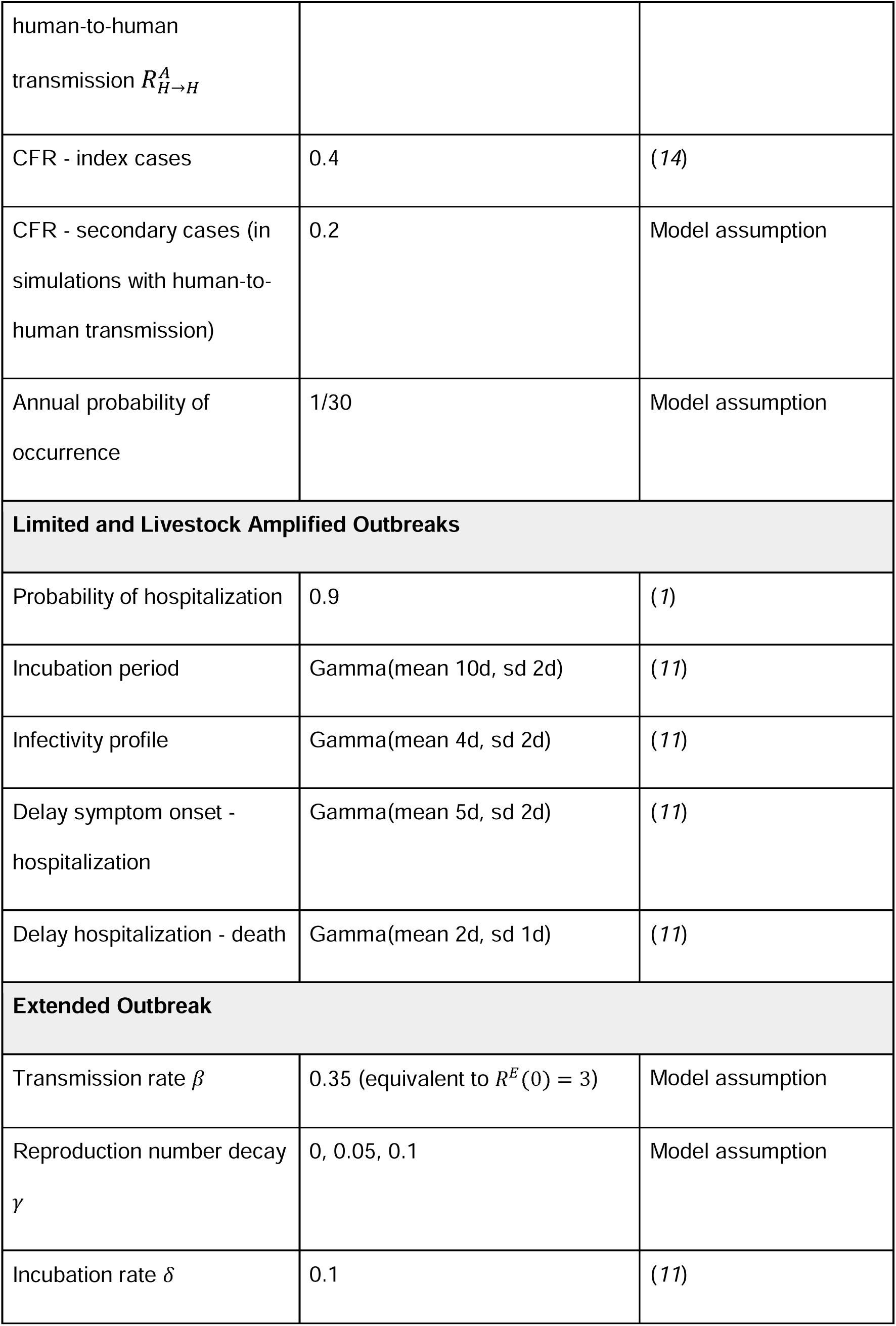
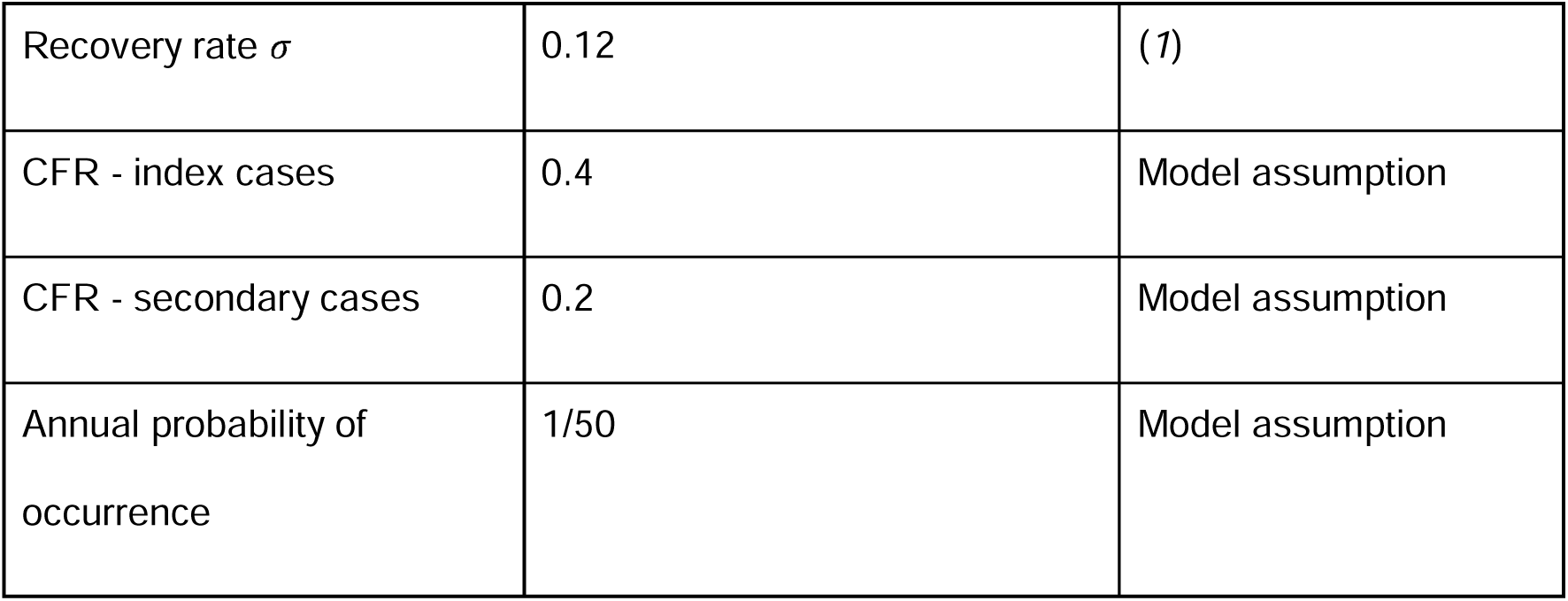
Model parameters of NiV transmission dynamics and natural history of disease.

### Livestock amplified outbreaks

In the livestock amplified scenario, we simulated outbreaks amongst livestock through a branching process model, where each case in livestock could also result in livestock-to-human transmission. For livestock-to-livestock transmission, a previous modelling study found that a basic reproduction number 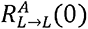 within the range 2.5 - 30 best reproduced the high levels of seroprevalence found in the pig farm where the 1998-1999 outbreak originated (*15*). We used the value 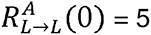, and we accounted for the natural depletion of susceptibles amongst livestock by drawing secondary cases from a Poisson distribution with a time-dependent effective reproduction number 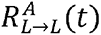 as follows:

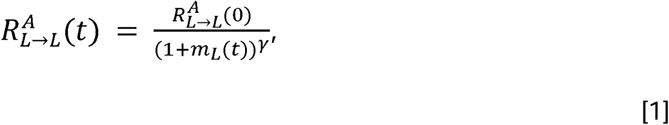

where *m_L_*(*t*) represents the number of infections accumulated in livestock at time t, and *γ*=0.2 informs the decay in the reproduction number (*22*). We simulated livestock-to-human transmission by drawing from a Poisson distribution with mean 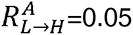, as inferred by a previous modelling study from case data from the 1998-1999 outbreak in Malaysia (*16*). Due to the lack of evidence of interhuman transmission during that outbreak, we did not consider human-to-human transmission in the base case of this scenario. Moreover, since there has been only one registered livestock amplified outbreak since 1998, we assumed that they occurred with an annual probability of 1/30. We conducted sensitivity analyses where interhuman transmission is possible, where each index human case triggered a limited outbreak simulation. In this scenario, index human cases resulted in death with a probability of 0.4, as observed during the 1998-1999 outbreak (*14*). In sensitivity analyses where human-to-human transmission was considered, we assumed that there would be a two-fold decrease in mortality between index and secondary cases, similarly to limited outbreaks. This resulted in a probability of mortality of 0.2 for secondary cases. All other parameters informing the natural history of disease and the delays between all stages of NiV transmission were identical to those used in limited outbreaks.

### Extended outbreaks

We modelled extended outbreaks through a stochastic, time-discrete SEIR compartmental model determined by:

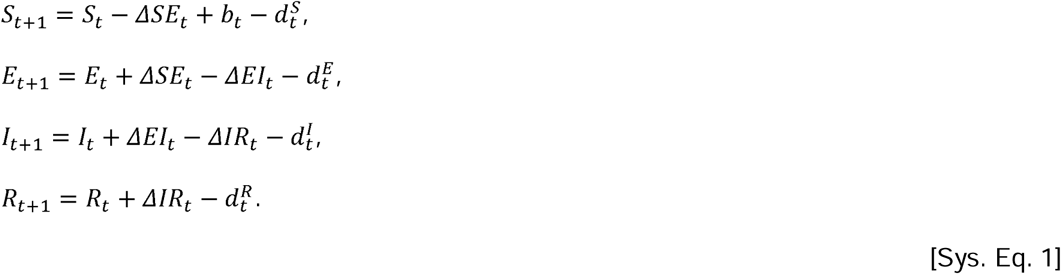

The transitions in the equation system are given by:

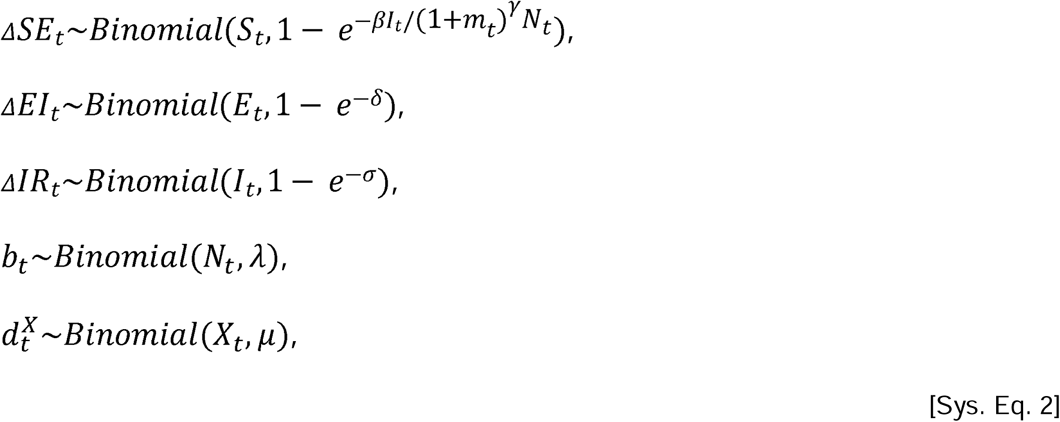

Where the transmission rate *β* is adjusted by the number of accumulated infections in the population at time t *m_t_*, and *γ* informs a decay in transmission intensity to reflect behavioral changes in the population to curb transmission throughout the outbreak. *N_t_ = S_t_ + E_t_ + I_t_ + R_t_* represents the total population size, *δ* = 0.1 represents the incubation rate (*11*), and *σ* = 0.12 represents the recovery rate (*1*). The parameters *λ* and *μ* represent the per capita birth and death rates, respectively.

We simulated extended outbreaks in a population with initial size 3.3 million, equivalent to the population of Faridpur and Raljbari districts, the two districts that most frequently report NiV outbreaks in Bangladesh (*4*), with one single exposed individual at the beginning of each simulation. We conducted simulations for 30 years with daily transitions, with an annual probability of outbreak occurrence of 1/50. Once an index case was generated, outbreaks unfolded with *β* = 0.35, equivalent to a *R^E^* (0) = 3, and we investigated the effect of different values for the decay in transmission intensity, with *γ* ∈ { 0, 0.05, 1 }.

For each outbreak scenario, we generated between 500 and 4,000 simulations, until we obtained at least 300 simulations where the number of cases matched the criteria for the corresponding scenario. We developed and implemented our models using R version 4.4.1 (*23*). Extended outbreaks were simulated using odin2 and dust2 (*24, 25*). All parameters used to simulate outbreaks in the base case and in sensitivity analyses are detailed in Table 1.

### Vaccine rollout and characteristics

Across all outbreak scenarios, we investigated the potential impact of different vaccine rollout strategies, logistical constraints, and product characteristics. We evaluated the impact of proactive and reactive vaccination strategies over a 30-year period, with a scenario-specific annual probability of outbreak occurrence. In proactive vaccine rollout simulations, we assumed that the population at risk was vaccinated at the beginning of the 30-year period, and that vaccination coverage would be maintained over the entire time period. In reactive vaccine rollout simulations, the hospitalization of the first case triggered a scenario-specific reactive campaign.

Reactive vaccination strategies were different for each outbreak scenario. In limited outbreaks, we simulated ring vaccination, where each hospitalization triggered vaccination in contacts and contacts of contacts. We assumed that each case had k = 17 contacts, and that 90% of them would be reached by contact-tracing efforts within five days (*1*). In livestock amplified outbreaks, we considered that individuals living within a 2km radius from infected farms would be targeted. This was a priority area for Japanese Encephalitis (JE) immunizations during the 1998-1999 outbreak in Malaysia (*26*), when NiV had not yet been identified as the causative agent. We assumed that 90% of individuals in the area would be vaccinated within 30 days. In extended outbreaks, we assumed that the general population would be targeted, with the same coverage and population at risk as with proactive vaccination.

For limited and livestock amplified outbreaks, we modeled vaccine distribution by randomly labelling individuals in simulated transmission chains according to the timing and coverage of the specific rollout strategy. For extended outbreaks, we added vaccinated compartments to our SEIR model (see Supplementary Information, Figure S6).

In proactive vaccination strategies, we assumed a vaccine coverage of 20%. In limited and extended outbreaks, we assumed that the population at risk that would be targeted by a vaccination campaign was the population of Faridpur or Rajbari districts, the two districts which have most frequently reported NiV outbreaks in Bangladesh (*4*). For livestock amplified outbreaks, we assumed that the population at risk would be the population of Perak and Negeri Sembilan states, the two states that reported the most number of cases during the 1998-1999 NiV outbreak in Malaysia (*14*).

As there are currently no licensed Nipah vaccines, we relied on target product characteristics, on assumptions from previous modeling studies, an expert panel with individuals from academia, and vaccine developers to inform parameters regarding key characteristics of a Nipah vaccine. We assumed that a single dose would provide lifelong immunity after 14 days (*11*). For limited and livestock amplified outbreaks, the vaccine provided protection against infection of 90% (*11*). In extended outbreaks, we explored the possibility of observing asymptomatic infections, and we distinguished between protection against infection of 70% and protection against disease of 90%. In reactive campaigns, we assumed that vaccine manufacture would take 75 days, and that their transport would require 2 days. In limited outbreak scenarios, where multiple outbreaks could take place over a 30-year period, we assumed that after the first outbreak, vaccines would be stockpiled and available after 2 days of transport.

We measured the potential impact of vaccines by counting how many cases, hospitalizations, deaths, and Disability-Adjusted Life Years (DALYs) would have been averted in each scenario and vaccination strategy. DALYs represent a measure of the burden of a disease, with one DALY representing the loss of one year of full health. We computed DALYs as follows:

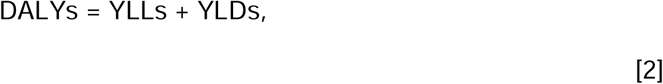

where YLLs represent years of life lost due to premature death, and YLDs represent years lived with a disability. For each simulated outbreak, we computed YLLs by multiplying the number of deaths by the difference between life expectancy and the scenario-specific mean age of cases. We computed YLDs by multiplying the number of cases who survive the acute phase of disease by the probability, average duration, and disability weight of different sequelae that have been reported in Nipah survivors (*13, 17, 27, 28*). The reported sequelae are different between survivors of outbreaks in Bangladesh and in Malaysia. We summarized their characteristics in Table S1. We note that studies that reported sequelae in NiV survivors were conducted at most 2 years after the initial phase of disease. Most reported sequelae were persistent at the time of the investigation. We assumed that all sequelae noted as persistent lasted for the rest of the individual’s life. We assumed that individuals infected in extended outbreaks would experience the same morbidity as individuals infected in livestock amplified outbreaks.

To account for uncertainty around transmission dynamics, vaccine characteristics, and feasibility of vaccination campaigns, we conducted sensitivity analyses where we relaxed simulation parameters one at a time. Parameters used in base case scenarios and in sensitivity analyses are summarized in Table 2.

**Table 2:** Vaccine product characteristics and logistical constraints.

| Parameter | Base case (sensitivity range) | Source |
| --- | --- | --- |
| **Vaccine Characteristics** |  |  |
| Vaccine efficacy (against disease in extended outbreaks) | 90% (Sensitivity: 75 - 100) | (11) |
| Vaccine efficacy against infection (extended outbreaks) | 70% (Sensitivity: 50 - 90) | Model assumption |
| Number of doses | Single dose (2 doses) | (11) |
| Delay to protection | 14 days (Sensitivity: 7-28 days) | Model assumption |
| Duration of protection | Life-long protection (Sensitivity: exponential decay with an average duration of 5 years) | (11, 30) |
| **Proactive Vaccination** |  |  |
| Target population | Limited/Extended outbreaks:<br>Rajbari and Faridpur districts, Bangladesh - 3.3 million<br>Livestock amplified outbreaks:<br>Perak and Negeri Sembilan states, Malaysia - 3.7 million | (4, 11, 14) |
| Coverage | 20% (Sensitivity: 10 - 40) | Model assumption |

| **Reactive Vaccination** |  |  |
| --- | --- | --- |
| Production delay | 75 days (Sensitivity: 46 - 152) | Pharmaceutical developer |
| Transport delay | 2 days | Model assumption |
| **Limited Outbreaks - Ring vaccination** |  |  |
| Target population | Contacts and contacts of contacts of hospitalized cases | Model assumption |
| Number of contacts of confirmed/suspect cases | 17 contacts | (1) |
| **Livestock Amplified Outbreaks - Spatial targeting of farms** |  |  |
| Target population | Population living within a 2km radius of farms | (26) |
| Vaccine rollout | 1 month (Sensitivity: 2 - 8 weeks) | Model assumption |
| **Extended Outbreaks - General population** |  |  |
| Target population | Rajbari and Faridpur districts, Bangladesh | (4, 11) |
| Vaccine rollout | 1 month (Sensitivity: 2 - 8 weeks) | Model assumption |

### Monoclonal antibody rollout and characteristics

We modeled the potential impact of monoclonal antibodies (mAbs) during reactive vaccination campaigns. According to target product characteristics of the MBP1F5 mAb, which is due to start Phase 2 clinical trials in Bangladesh and India in 2026, we incorporated mAbs used as Pre-Exposure Prophylaxis (PrEP), Post-Exposure Prophylaxis (PEP), or as therapeutics. We assumed that 10,000 mAb doses would be stockpiled and would be distributed after two days of lead transport time.

We assumed that PrEP would be delivered to individuals at high risk of NiV exposure during an outbreak, prioritizing healthcare workers (HCWs). In limited outbreaks, we modeled PrEP delivery by randomly selecting 30% of cases, and by delivering PrEP if they had not been vaccinated and had not experienced symptoms at the time. Since livestock amplified outbreaks would primarily affect farmers, in this scenario PrEP was only administered to 30% of secondary cases in sensitivity analyses with human-to-human transmission. In extended outbreaks, we added PrEP compartments to our SEIRV model, with similar characteristics as those detailed in equation systems 3, and 4. We assumed that all 10,000 doses would be delivered within a period of 30 days (Figure S6). We evaluated if this would be sufficient to target HCWs in Faridpur and Rajbari districts in Bangladesh assuming that publicly employed physicians, nurses, community medical officers, medical technologists, and hospital domiciliary staff would be targeted. For the first four categories, a report from the Bangladeshi Ministry of Health and Family Welfare (MoHFW) estimates 6.54 HCWs per 10,000 population in Dhaka division, which includes Rajbari and Faridpur districts (*18*). For domiciliary staff, we assumed that the spatial distribution of these HCWs across divisions was the same as for the rest, and we used a separate estimate from the MoHFW of 57,000 domiciliary staff employed in Bangladesh (*19*).

For mAbs used as PEP, in limited outbreaks and in livestock amplified outbreaks with human-to-human transmission, we considered that the hospitalization of a case would trigger PEP delivery to 90% of their unvaccinated contacts within 5 days if the contacts had not declared symptoms. In this case, only contacts of contacts would receive vaccination. In extended outbreaks, we added PEP compartments to our SEIRV model (Figure S6). The proportion of contacts of hospitalized patients receiving PEP that were exposed to NiV was given by a parameter a. In the context of observed NiV outbreaks in Bangladesh, it is estimated that 1.4% of contacts of NiV cases result in human-to-human transmission (*1*). We assumed that, in the extended scenario, the increase in this parameter would be proportional to the increase in the basic reproduction number, leading to a value of *α* = 12.7%.

Finally, for the therapeutic use of mAbs, we assumed that these would be delivered to all individuals with encephalitis symptoms upon hospitalization, regardless of vaccination status, within the limit of 10,000 available doses. We accounted for the fact that mAbs would be used on patients that were not infected with NiV using previous studies that estimate that 2% of encephalitis patients test positive for NiV in surveillance hospitals (*4*). We scaled this proportion with the reproductive number of the outbreak scenario. For the simultaneous use of mAbs as PrEP, PEP, and therapeutics, we considered that individuals could only receive mAbs under one of these pathways.

In our baseline case, PrEP and PEP had an efficacy of 95% to prevent disease and onwards NiV transmission. In our model, the therapeutic use of mAbs could only prevent death, with an efficacy of 95% (*29*). We assumed that the passive immunity provided by PrEP and PEP would have a duration of 6 months maximum. We conducted sensitivity analyses around the characteristics of mAbs, which are detailed in Table 3. We incorporated mAb use in reactive vaccination strategies for all simulated outbreak scenarios, and we measured their impact in terms of cases, hospitalizations, deaths, and DALYs averted.

**Table 3:** Monoclonal antibody product characteristics and logistical constraints.

| Parameter | Base case (sensitivity range) | Source |
| --- | --- | --- |
| **mAb characteristics** |  |  |
| Efficacy against disease and onwards transmission (PrEP and PEP) | 95% (Sensitivity: 50 - 100) | Model assumption based on (29) |
| Efficacy against death (Therapeutic) | 95% (Sensitivity: 50 - 100) | (29) |
| Number of doses | Single dose (Sensitivity: 2 doses for PEP/Therapeutic) | Target product characteristic |
| Duration of protection | 6 months (Sensitivity: 3 - 12) | Model assumption |
| **Rollout** |  |  |
| Target Population: PrEP | Healthcare workers | (18, 19) |
| Target Population: PEP | Individuals with a known or suspected exposure to NiV prior to symptom onset (contacts of hospitalized NiV cases) | Model assumption |
| Target Population: Therapeutic | Encephalitis cases upon hospitalization | Model assumption |
| Production delay | 90 days (Sensitivity: 0 - 120) | Pharmaceutical developer |
| Transport delay | 2 days | Model assumption |
| Stock | 10,000 doses | Model assumption |

## Results

We simulated NiV outbreaks over a 30-year period under limited, livestock amplified, and extended outbreak scenarios. Limited outbreaks were simulated with a branching process model informed by surveillance data from NiV outbreaks in Bangladesh (Table 1, Figure 1) (*1, 11, 12*). Individual outbreaks had a mean size of 5.8 cases (95%CI: 2-26), 5.3 hospitalizations (95%CI: 1-25), and 3.1 deaths (95%CI: 1-13) (Figure 2A). Using follow-up data on long-term sequelae among NiV survivors in Bangladesh, these infections resulted in 174 DALYs (95%CI: 57-748) per outbreak (parameters are detailed in Table S1) (*13*).

**Figure 1:**
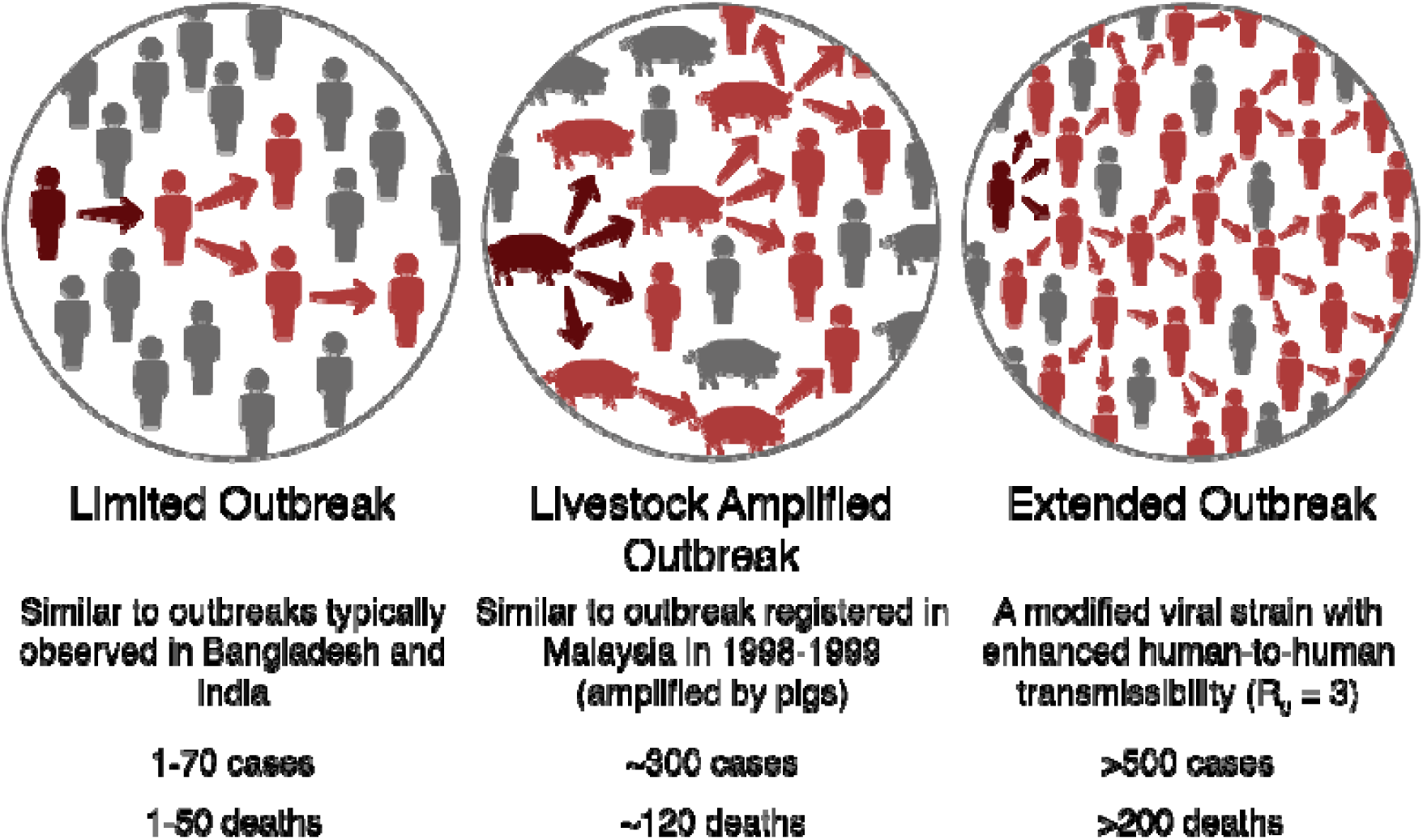
Schematic representation of the three simulated outbreak scenarios, with key metrics.

**Figure 2:**
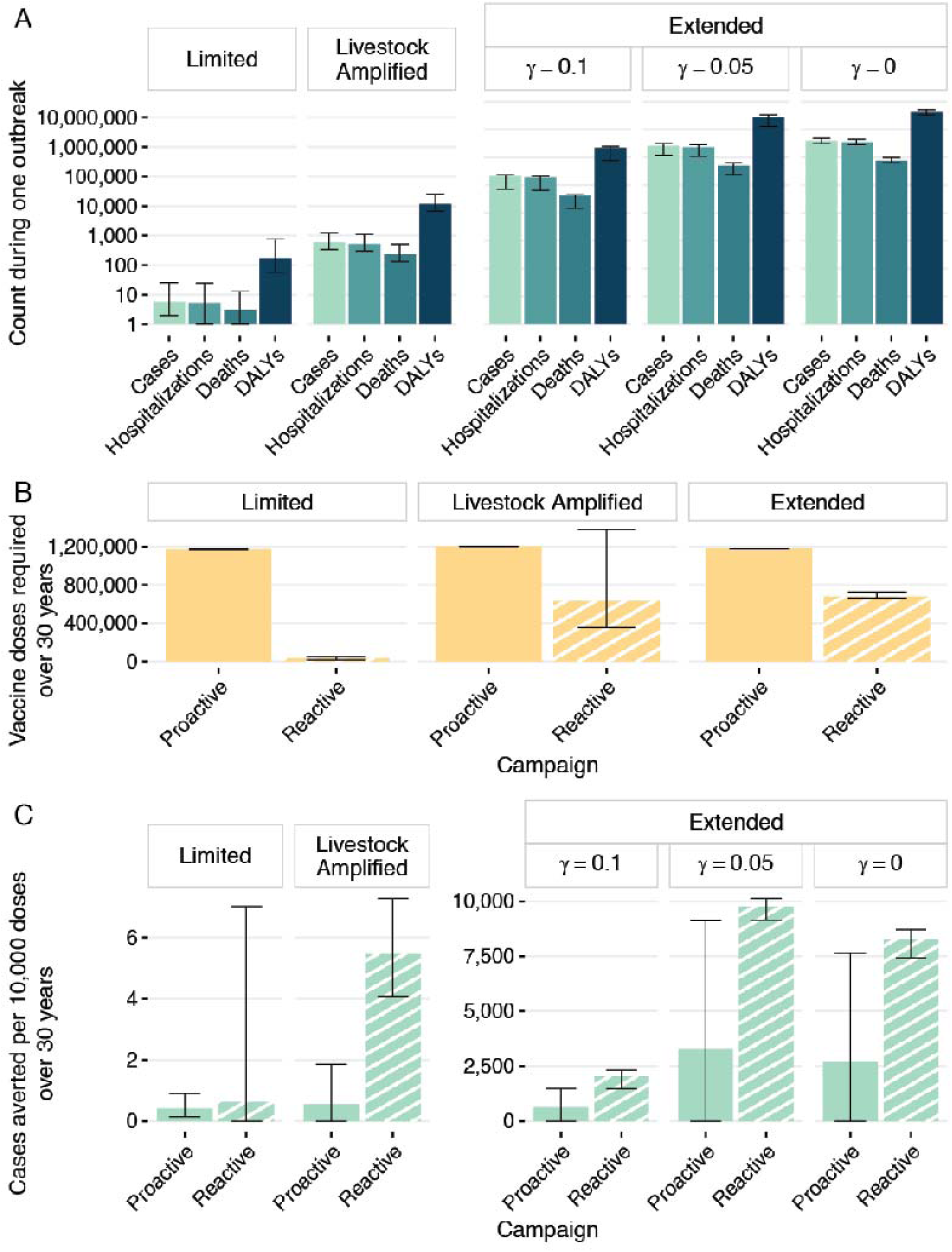
**(A)** Estimated burden for one outbreak in each epidemiological scenario, measured in cases, hospitalizations, deaths, and DALYs; **(B)** Number of doses required over 30 years for proactive and reactive vaccination in each epidemiological scenario; **(C)** Averted cases per 10,000 vaccine doses over 30 years for each epidemiological scenario, with proactive and reactive vaccination strategies.

In livestock amplified outbreaks, we simulated NiV transmission in livestock using a branching process model informed by the 1998-1999 outbreak across pig farms in Malaysia (Table 1, Figure 1) (*14, 15*). Humans could then be infected by livestock (*16*). Due to a lack of evidence of human-to-human transmission during the 1998-1999 outbreak in Malaysia, we did not incorporate this transmission pathway in our base case for this outbreak scenario. Under these assumptions, we estimate 386 cases (95%CI: 333-447), 348 hospitalizations (95%CI: 298-404), and 155 deaths (95%CI: 128-185) per outbreak (Figure 2A). Moreover, using information from the follow-up of NiV survivors from the 1998-1999 outbreak in Malaysia, we estimate that these infections would result in 7,800 DALYs (95%CI: 6,500- 9,300) per outbreak (parameters are detailed in Table S1) (*17*). Since only one outbreak of the kind has been detected since NiV was first isolated, we assumed that livestock amplified outbreaks had an annual probability of occurrence of 1/30.

We next considered the possible emergence of a more transmissible NiV strain, leading to widespread or extended outbreaks, assuming that this strain would have the same morbidity and mortality as observed during the 1998-1999 outbreak in Malaysia. We simulated these with a time-discrete, stochastic SEIR model. We considered a population the size of Rajbari and Faridpur districts in Bangladesh, since these are the districts that report outbreaks most frequently, and we implemented the model using a SARS-like basic reproduction number of 3 (Table 1, Figure 1). In an outbreak resulting in thousands of Nipah cases, it is likely that there would be behavioral changes in the population, leading to a decay in viral transmission. We investigated the effects of three different levels of transmission dampening: no decay, intermediate, and high (Figure 2A, Figure S1). With high decay in transmission, a single extended outbreak resulted in 206,000 cases (95%CI: 198,000- 217,000). We assumed that the NiV strain would have comparable mortality and morbidity levels to those observed during the 1998-1999 outbreak in Malaysia, leading to 186,200 hospitalizations (95%CI: 178,000-195,000), 41,000 deaths (95%CI: 40,000-43,000), and 2.2 million DALYs (95%CI: 2.1-2.3) per outbreak. Intermediate transmission decay resulted in 10.4 times more cases (95%CI: 9.9-10.9), and no decay resulted in 15.3 times more cases (95%CI: 14.6-16). Additionally, we assumed that these outbreaks could occur with an annual probability of 1/50 to denote their lower likelihood compared to limited and livestock amplified scenarios.

In limited and livestock amplified outbreaks, we evaluated the potential impact of a single-dose vaccine with an efficacy against infection and disease of 90%, due to the lack of evidence for asymptomatic infections with NiV strains observed so far. In extended outbreaks, we investigated the possibility of a vaccine that would confer 70% protection against infection, and 90% protection against disease. We overlaid proactive and scenario-specific reactive immunization strategies over simulated outbreaks. For the reactive campaigns we assumed the vaccine only became available after 75 days of lead manufacture time.

In the limited scenario, we incorporated mass proactive vaccination of 20% of the population of Rajbari and Faridpur districts. As a reactive vaccination strategy, we considered the effects of ring vaccination, where contacts and contacts of contacts of hospitalized patients were vaccinated. Over a 30-year period, proactive vaccination required 1.2 million doses and averted 48 cases (95%CI: 17-105), 44 hospitalizations (95%CI: 16-94), 24 deaths (95%CI: 9-50), and 1,400 DALYs (95%CI: 500-2,900) (Figure 2A). In reactive vaccination, 31,000 doses were used (95%CI: 16,000-52,000), and 2 cases (95%CI: 0-27), 2 hospitalizations (95%CI: 0-24), 1 death (95%CI: 0-10), and 60 DALYs (95%CI: 0-630) were averted (Figure 2B). The impact of both immunization strategies was comparable when measured in the number of cases averted per 10,000 doses over 30 years, estimated at 0.41 (95%CI: 0.15- 0.9) for proactive vaccination and at 0.62 (95%CI: 0-7) for reactive vaccination (Figures 2C, S2). Reducing the lead time from detection of a hospitalized case to reactive vaccination (i.e., assuming there is an accessible stockpile at all times), would result in 0.64 cases (95%CI: 0-7.1) averted per 10,000 doses over 30 years and require an average of 1,100 doses (95%CI: 600-1,800) per year.

For proactive vaccination in livestock amplified outbreaks, we considered that 20% of the population of Perak and Negeri Sembilan states in Malaysia, the states that registered most cases during 1998-1999, would receive vaccination. Over 30 years, this approach averted a mean of 67 cases (95%CI: 0-223), 60 hospitalizations (95%CI: 0-200), 27 deaths (95%CI: 0- 93), and 1,350 DALYs (95%CI: 0-4,680) with 1.2 million vaccine doses used. In reactive vaccination, we assumed that individuals within a 2km radius of affected farms would be targeted and resulted in a mean of 344 cases (95%CI: 182-773), 309 hospitalizations (95%CI: 164-689), 137 deaths (95%CI: 70-311) and 6,900 DALYs averted (95%CI: 3,500-15,800) with 630,000 vaccine doses (95%CI: 360,000-1,400,000) used. When considering the impact per dose, we find that reactive vaccination would be most effective, averting 5.4 cases (95%CI: 4-7.2) per 10,000 doses over 30 years, compared to 0.6 cases (95%CI: 0- 1.8) per 10,000 doses over 30 years with proactive vaccination (Figures 2C, S2).

In extended outbreaks, both for proactive and reactive vaccination, we assumed that 20% of the general population of Rajbari and Faridpur districts in Bangladesh was vaccinated. Over 30 years, proactive vaccination required 1.2 million doses (Figure 2B). With high transmission decay, this rollout strategy averted an average 73,000 cases (95%CI: 0- 174,000), 65,000 hospitalizations (95%CI: 0-157,000), 15,000 deaths (95%CI: 0-35,000), and 0.77 million DALYs (95%CI: 0-1.9) over 30 years. Reactive vaccination resulted in 141,000 cases (95%CI: 101,000-159,000), 127,000 hospitalizations (95%CI: 91,000-143,000), 28,000 deaths (95%CI: 20,000-32,000), and 1.5 million DALYs averted (95%CI: 1.1-1.7), requiring 690,000 doses (95%CI: 674,000-733,000). Adjusting for vaccine doses, with reactive vaccination, there were 2,000 cases averted per 10,000 doses (95%CI: 1,500- 2,300), compared to 600 (95%CI: 0-1,500) with proactive vaccination (Figures 2C, S2). Similarly, reactive vaccination was more effective with intermediate and with no transmission decay (Figure 2C).

Monoclonal antibodies (mAbs) can provide immediate passive immunity and can therefore be a useful tool to mitigate viral spread. We included the use of mAbs as pre-exposure prophylaxis (PrEP) prioritizing healthcare workers (HCWs), as post-exposure prophylaxis (PEP) for contacts of hospitalized Nipah patients, or as therapy for hospitalized cases, in addition to reactive vaccination for all outbreak scenarios. Across all outbreak scenarios, we assumed that a stockpile of 10,000 mAb regimens would be available during an outbreak after 2 days of transport lead time. In limited outbreaks, PrEP increased the average number of DALYs averted over 30 years to 480 (95%CI: 56-1,400), PEP increased them to 730 (95%CI: 39-2,200). Therapeutic mAbs had the strongest effect, averting 2,000 DALYs over the same time (95%CI: 950-3,600). In livestock amplified outbreaks, since we do not include human-to-human transmission in our base case scenario, no PrEP or PEP regimens were used (Figure 3). The use of all 10,000 therapeutic doses raised the number of averted DALYs to 11,000 (95%CI: 6,500-24,000) (Figure 3B). Across all levels of transmission decay in extended outbreaks, PrEP and PEP had comparable effects: 1.7 million (95%CI: 0.69-1.9) and 1.7 million (95%CI: 0.68-1.9) DALYs averted in outbreaks with high transmission decay; 8.9 million (95%CI: 7.2-12.1) and 8.9 million (95%CI: 7.2-12) DALYs averted with intermediate decay; and in extended outbreaks with no transmission decay, both PrEP and PEP averted 7.3 million DALYs (95%CI: 5.9-9). Therapeutic mAbs also had comparable effects, though marginally stronger in high transmission decay outbreaks, with 1.9 million DALYs averted (95%CI: 1.5-2.2), and marginally weaker in intermediate and no transmission decay outbreaks, with 7.8 million (95%CI: 7.7-8.2) and 6.9 million (95%CI: 6.6-7.3) DALYs averted, respectively (Figure 3). Additionally, we estimated that targeting HCW across Rajbari and Faridpur districts in Bangladesh would require ∼3,000 PrEP doses (*18, 19*).

**Figure 3:**
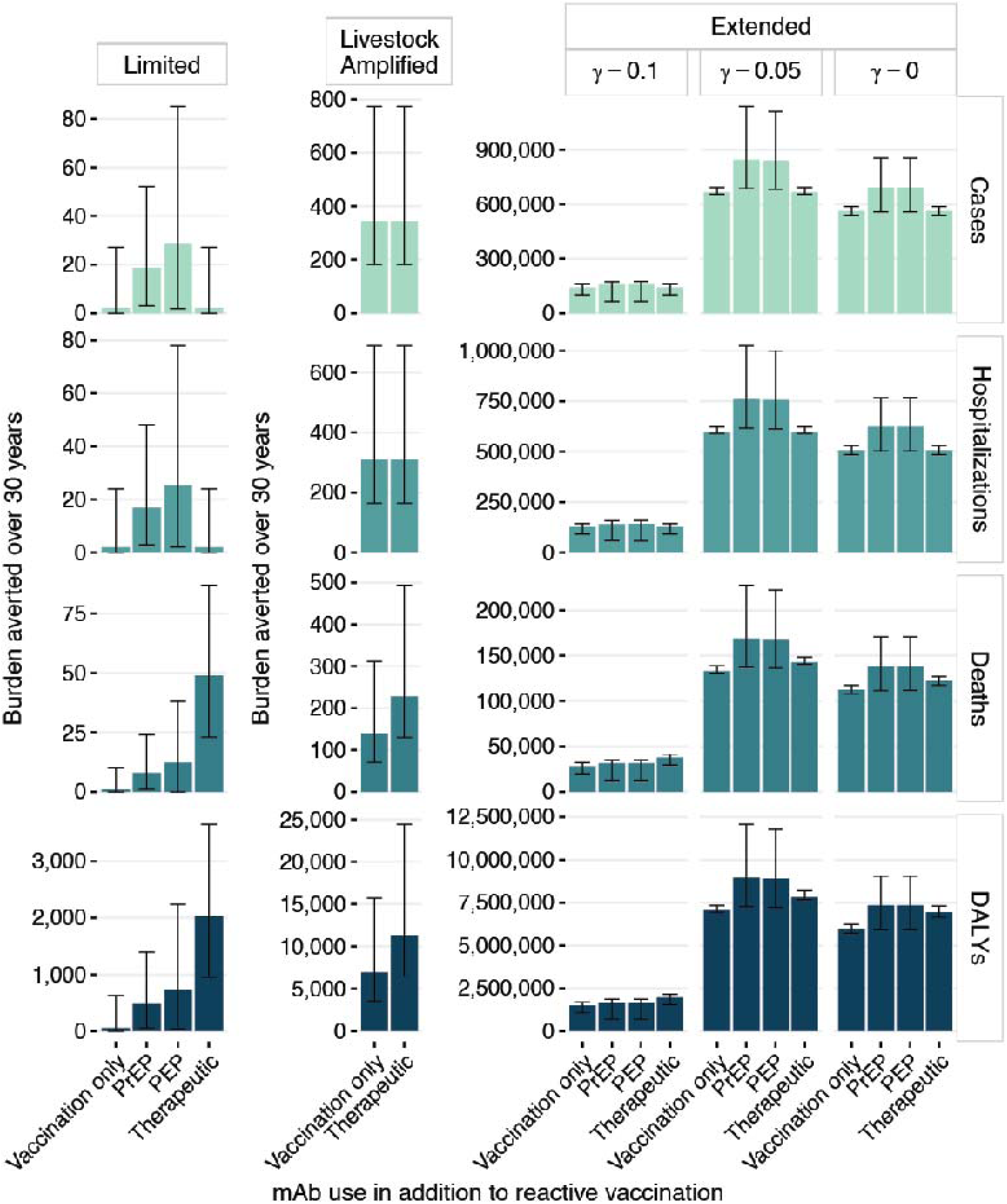
Burden averted when 10,000 mAb doses are used as PrEP, as PEP, or as therapy in addition to reactive vaccination. Bars represent mean estimates and error bars represent 95% confidence intervals.

To account for underlying uncertainty in future transmission dynamics, in vaccine characteristics, and in the feasibility and reach of rollout campaigns, we conducted sensitivity analyses where we allowed one parameter to vary at a time (see Methods and Tables 1, 3, 4). We found that across all outbreak scenarios, the potential impact of reactive campaigns were most sensitive to variations in parameters determining transmission dynamics, such as the basic reproduction number in limited outbreaks (Figure 4A). In livestock amplified outbreaks, where we considered interhuman transmission as a sensitivity, this was the case when varying the basic reproduction number both in livestock-to-human and in human-to-human transmission (Figure 4B). In extended outbreaks, the level of transmission decay as infections accumulated, and the basic reproduction number in outbreaks with high transmission decay had the strongest effects (Figures 2, 4). In limited outbreaks, lag time to protective immunity, vaccine efficacy, the number of doses of the vaccine, and vaccine coverage also had a substantial effect on averted burden. Livestock amplified outbreaks were also sensitive to vaccine efficacy, lag time to vaccine distribution, and vaccine coverage. Finally, in extended outbreaks with high transmission decay, vaccine coverage and vaccine efficacy against infection also led to changes in the levels of burden averted. Other levels of transmission decay were less sensitive to variations in the basic reproduction number (Figure S4). Patterns of sensitivity of vaccine impact were consistent when considering proactive vaccination (Figure S5). In sensitivity analyses of mAb distribution, all different mAb uses were sensitive to changes in lead time to mAb distribution and to mAb efficacy (Figure S3). PrEP use was also sensitive to the time over which mAbs were distributed.

**Figure 4:**
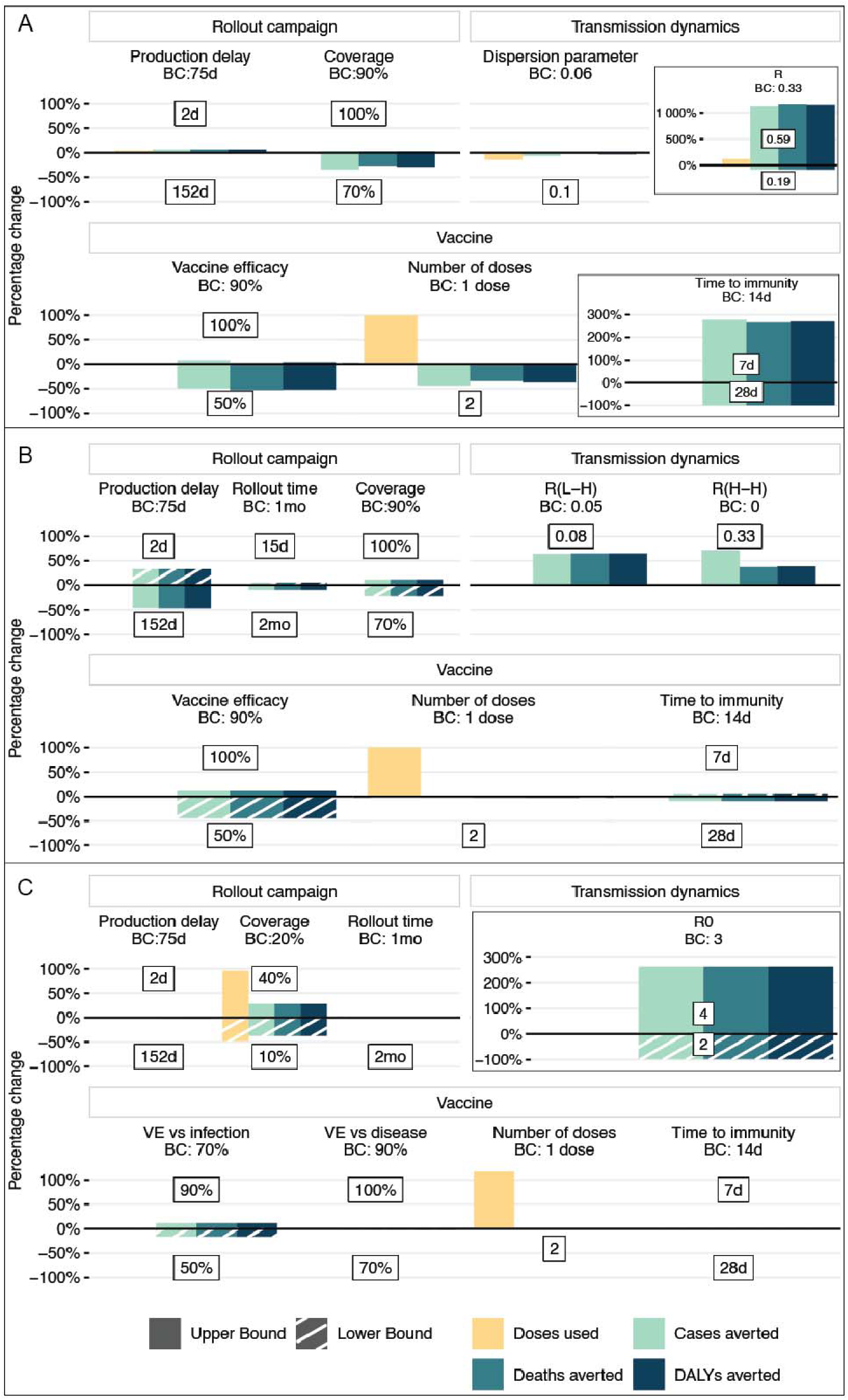
Sensitivity analysis of differences in potential vaccine impact when one parameter changes in comparison to the base case (BC) scenario for reactive vaccination in **(A)** limited, **(C)** livestock amplified, and **(C)** extended outbreaks with a high level of transmission decay due to behavioral changes (*γ* = 0.1). Comparisons are represented as the percentage change of median estimates of doses used, and cases, deaths, and DALYs averted.

## Discussion

Here we simulated NiV outbreaks according to two previously observed epidemiological scenarios (limited and livestock amplified) and to a third potential scenario in which the emergence of NiV strains with enhanced transmissibility would lead to widespread outbreaks. For the two previously observed types of outbreaks, we used parameters informed by surveillance and historical outbreak data, and our estimates matched burden levels that have been observed during real NiV outbreaks. We evaluated and compared the potential impact of medical countermeasures - vaccines and mAbs - for different deployment strategies.

Limited outbreaks are currently seen annually in Bangladesh and have been registered multiple times in India, and represent the scenario where vaccines may be most immediately used. However, we found that ring vaccination - targeting contacts and contacts-of-contacts of hospitalized Nipah patients, would have limited impact in this scenario, because most transmission events take place before hospitalization of the infector. Proactive vaccination would avert more cases but require many more doses, since the unpredictability of NiV outbreaks and the lack of clearly defined at-risk groups mean that large administrative regions would need to be targeted. Consequently, when evaluating these two immunization strategies by cases averted per 10,000 doses, they had a comparable effect. In the absence of a stockpile, long lead manufacture times would also mean that the impact of mAbs on cases averted would be negligible in limited outbreaks. Ultimately, for the most effective use of medical countermeasures in this scenario, there needs to be a stockpile of accessible vaccines and mAbs available during the NiV season. For the two most affected regions of Bangladesh, this would require an average stockpile of 1,100 vaccine doses. A stockpile of 10,000 mAb doses would be sufficient to target HCWs with PrEP (needing ∼3,000 doses, see Methods), allowing for the use of mAbs as PEP and therapy during outbreaks. The feasibility of establishing such a stockpile will depend on shelf life, production capacity of the vaccine and mAbs and logistical constraints in delivering them in a timely manner to rural regions.

In livestock amplified and extended outbreaks, reactive vaccination strategies - spatially targeting individuals near affected farms in the former and targeting the general population in the latter - outperformed proactive vaccination both in terms of overall burden averted, as well as when adjusting averted burden by the number of doses required. Even though proactive vaccination has the advantage of ensuring that the target population is protected before an outbreak is declared, its impact is reduced by the low annual probability of occurrence of these outbreaks in our simulation framework. This means that there were many 30-year simulations in which no outbreaks occurred, so no cases were averted while over 1 million vaccine doses were used each time. We note that livestock amplified outbreaks were noticeably sensitive to shorter lead manufacture times. Reducing the time to vaccine distribution from 77 to 2 days led to 33% more cases reduced on average, while an increase to 152 days resulted in 46% fewer cases averted. Hence, the ability to deliver vaccines quickly in this type of scenario could substantially increase vaccine impact. Furthermore, results from our simulation framework suggest that even though mAbs could be a useful tool to mitigate NiV outbreaks across all scenarios, the speed at which they would become available in limited outbreaks is also critical in ensuring their impact. Increasing the lead time to mAb distribution from 2 to 92 days resulted in a reduction in averted DALYs of 88% for PrEP, 91% for PEP, and 97% for therapeutic mAbs.

This study is subject to some limitations. Critically, NiV remains a poorly understood pathogen, and as for many zoonoses, the probability of spillover is determined by the interplay of a wide range of factors, like viral shedding in bats, pathogen survival, human behavior, and agricultural practices (*20*). For instance, spillover pathways for several NiV outbreaks in Kerala, India, between 2018 and 2024, remain unclear (*2*). Additionally, we based our simulation of livestock amplified outbreaks on the only outbreak of the kind that has been observed so far. This assumption allowed us to base our simulation on observed data, however, the size of potential future livestock amplified outbreaks could vary due to differences in agricultural practices and epidemic preparedness across countries and subnational regions where *Pteropus* bats circulate. Finally, there have been few studies describing long-term sequelae of Nipah disease, and survivors have been followed up for at most two years after outbreaks, which adds uncertainty to our DALY estimates (*13, 17*). Nonetheless, in our study DALYs are a linear combination of cases and deaths, so conclusions around what rollout strategies would avert most DALYs are robust to this uncertainty.

Vaccines seem to have restricted impact in mitigating burden during limited outbreaks, the most frequently observed scenario. Using stockpiled mAbs as therapeutics during these outbreaks could increase the impact of medical countermeasures, however, this hinges on high mAb efficacy and on product availability. In our simulations, therapeutic mAbs were more impactful than PrEP and PEP during limited outbreaks because they prevented death in individuals that had already developed disease. Similarly to vaccines, PrEP and, to a lesser extent, PEP had a reduced impact because most transmission events had taken place before the infector was hospitalized and the outbreak was detected. In livestock amplified outbreaks, vaccines were more impactful, since there were more generations in transmission chains. Here again, therapeutic mAbs increased the burden averted during outbreaks. Vaccines could be the most impactful during extended outbreaks, however, this impact relies on the distribution of many vaccine doses during one single outbreak. In outbreaks with the highest burden levels (scenarios with intermediate or no transmission decay as cases accumulated), the use of stockpiled mAbs as PrEP and PEP led to some increase in burden reduction.

*Pteropus* bats circulate over extensive areas in South and Southeast Asia, and previous studies suggest that up to 90% of NiV’s genetic diversity remains unobserved (*21*). In consequence, outbreaks in areas that have rarely or that have never experienced NiV, as well as the emergence of more transmissible viral strains, are plausible. Our study investigated the potential impact of vaccines under different epidemiological scenarios, filling a critical knowledge gap in epidemic and pandemic preparedness. Continued efforts in the development of medical countermeasures to ensure their timely distribution will be critical to mitigate NiV spread during outbreaks.

## Funding

OCA and HS acknowledge funding from the Coalition for Epidemic Preparedness Innovations. SC acknowledges support from the Investissement d’Avenir program, the Laboratoire d’Excellence Integrative Biology of Emerging Infectious Diseases program (Grant ANR-10-LABX-62-IBEID), the European Commission under the EU4Health programme 2021-2027, Grant Agreement - Project: 101102733 - DURABLE and the INCEPTION project (PIA/ANR-16-CONV-0005).

### Ethics statement

This work was conducted using simulated data and therefore did not require ethical approval.

### Data and code availability

All parameter values and code necessary to reproduce these analyses is available in the following GitHub repository: https://github.com/ocortesazuero/NiV_Vaccine_Impact.

## Data Availability

This work was conducted using simulated data and therefore did not require ethical approval. All parameter values and code necessary to reproduce these analyses is available in the following GitHub repository: https://github.com/ocortesazuero/NiV_Vaccine_Impact.

## Supplementary Information

### Vaccine impact in extended outbreaks

In extended outbreaks, we modified our SEIR model to add vaccinated compartments as follows:

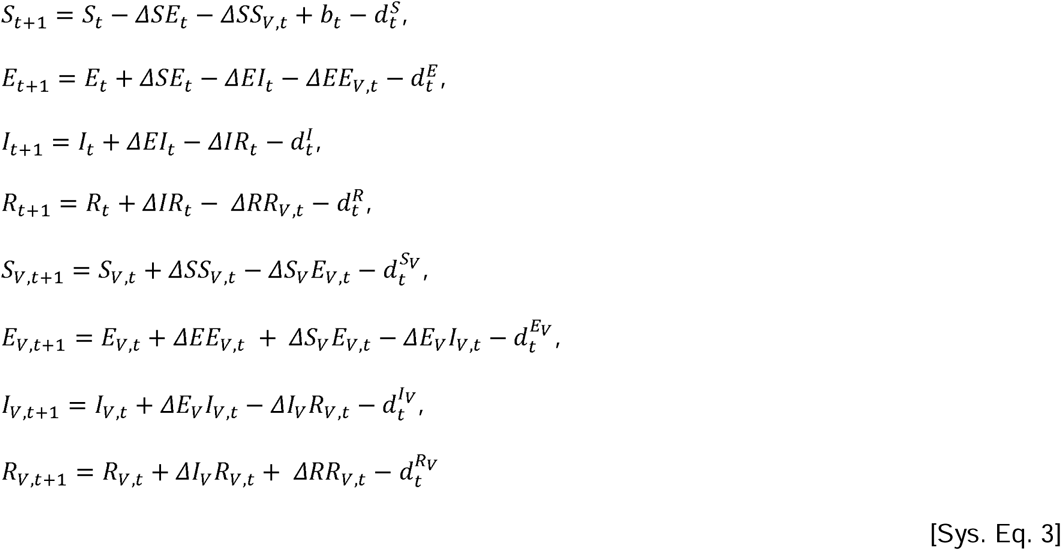

The transitions in the equation system are given by:

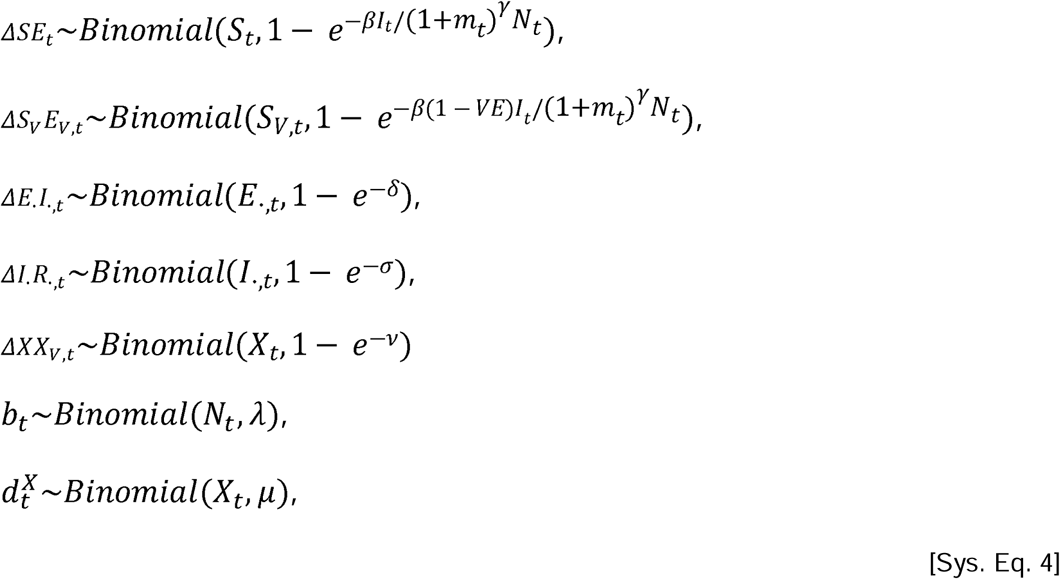

Where, in addition to parameters explained in equation systems 1 and 2, *VE* = 90% represents vaccine efficacy, and *v* represents the vaccination rate, equivalent to vaccinating 20% of the population in 30 days.

---

**Figure S1:**
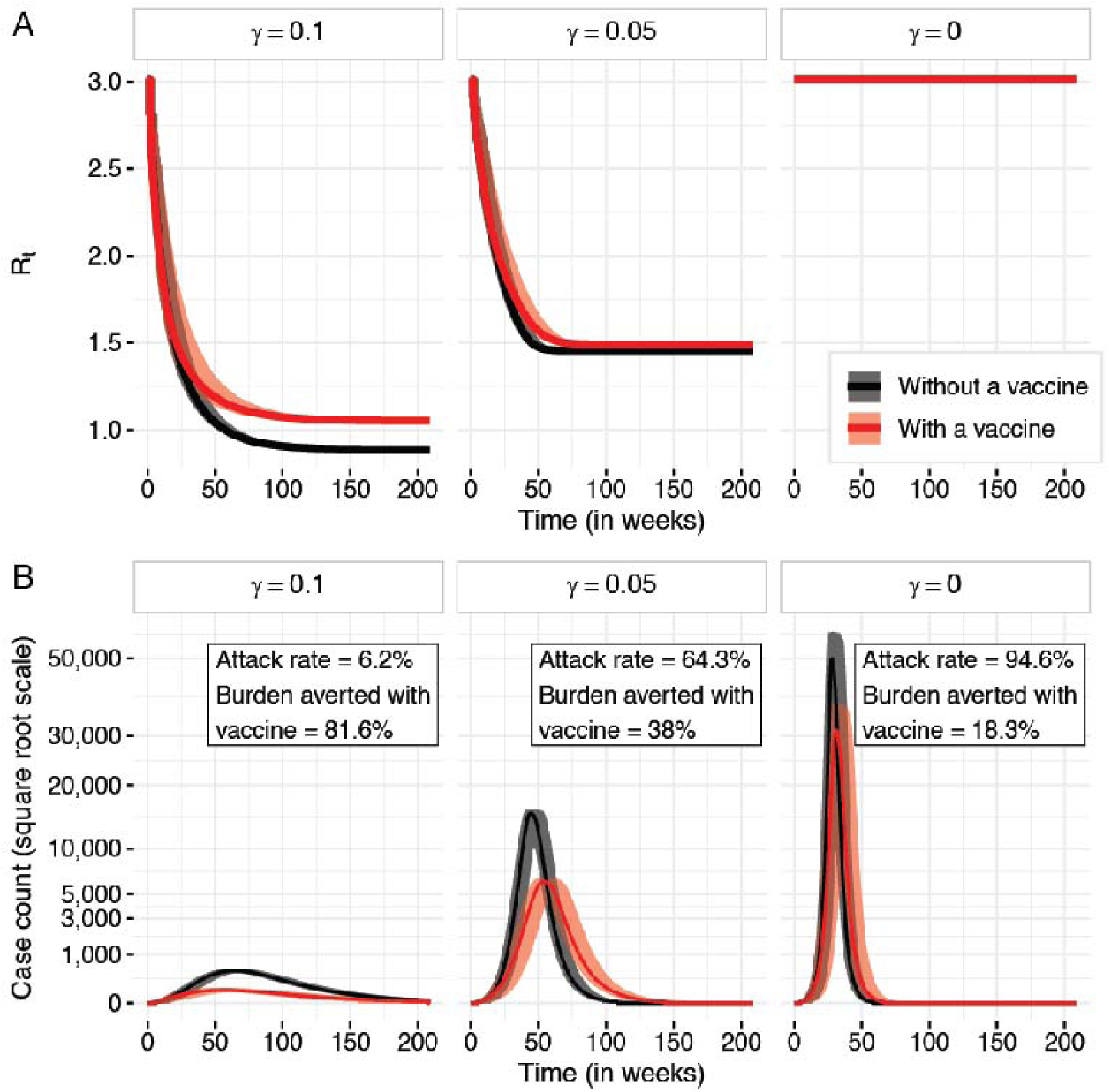
**(A)** Evolution of reproduction number over time and **(B)** epidemic curve, with and without vaccination, according to different levels of transmission decay due to behavioral changes in the population. Lines represent median values and ribbons represent 95% confidence intervals.

**Figure S2:**
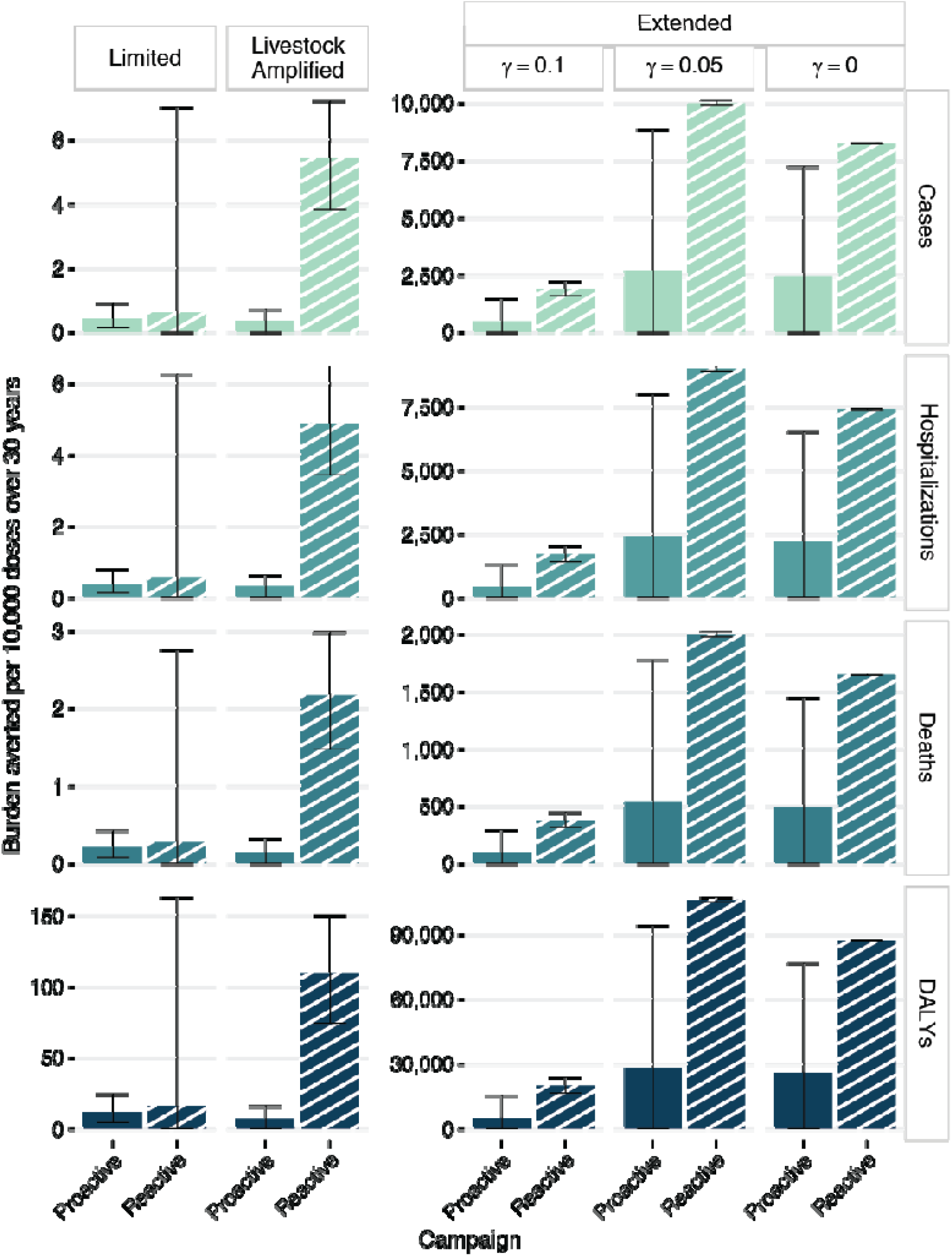
Burden averted per 10,000 vaccine doses used over 30-year simulations in terms of cases, hospitalizations, deaths, and DALYs for the three types of outbreak scenarios, with proactive and reactive vaccination campaigns. Bars represent mean estimates, and error bars represent 95% confidence intervals.

**Figure S3:**
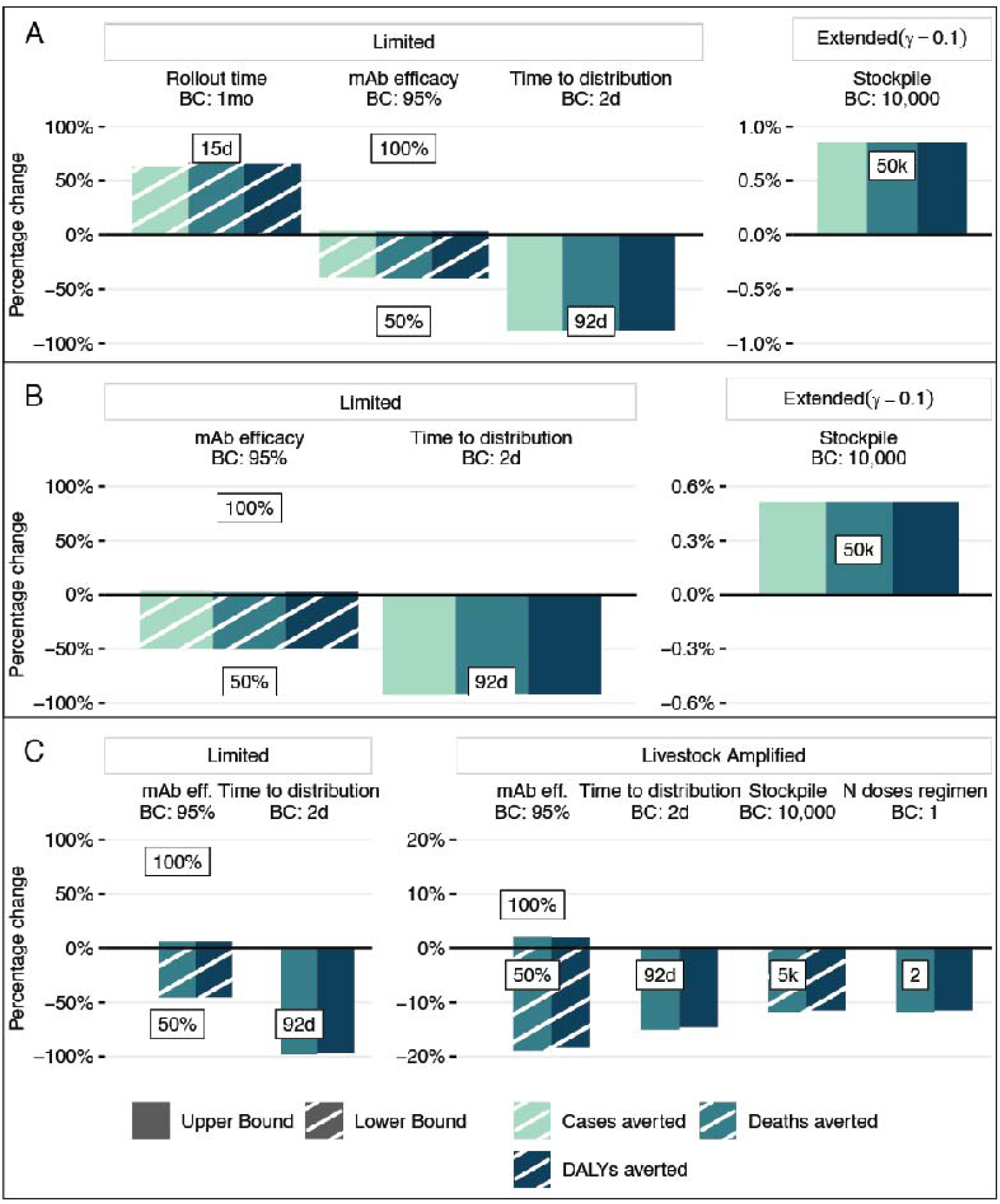
Sensitivity analysis of differences in potential impact when one parameter changes in comparison to the base case (BC) scenario for reactive vaccination with mAbs used as **(A)** PrEP, **(B)** PEP, **(C)** or therapy. Comparisons are represented as the percentage change of median estimates of cases, deaths, and DALYs averted. Only parameters with a variation of more than 0.5% are displayed.

**Figure S4:**
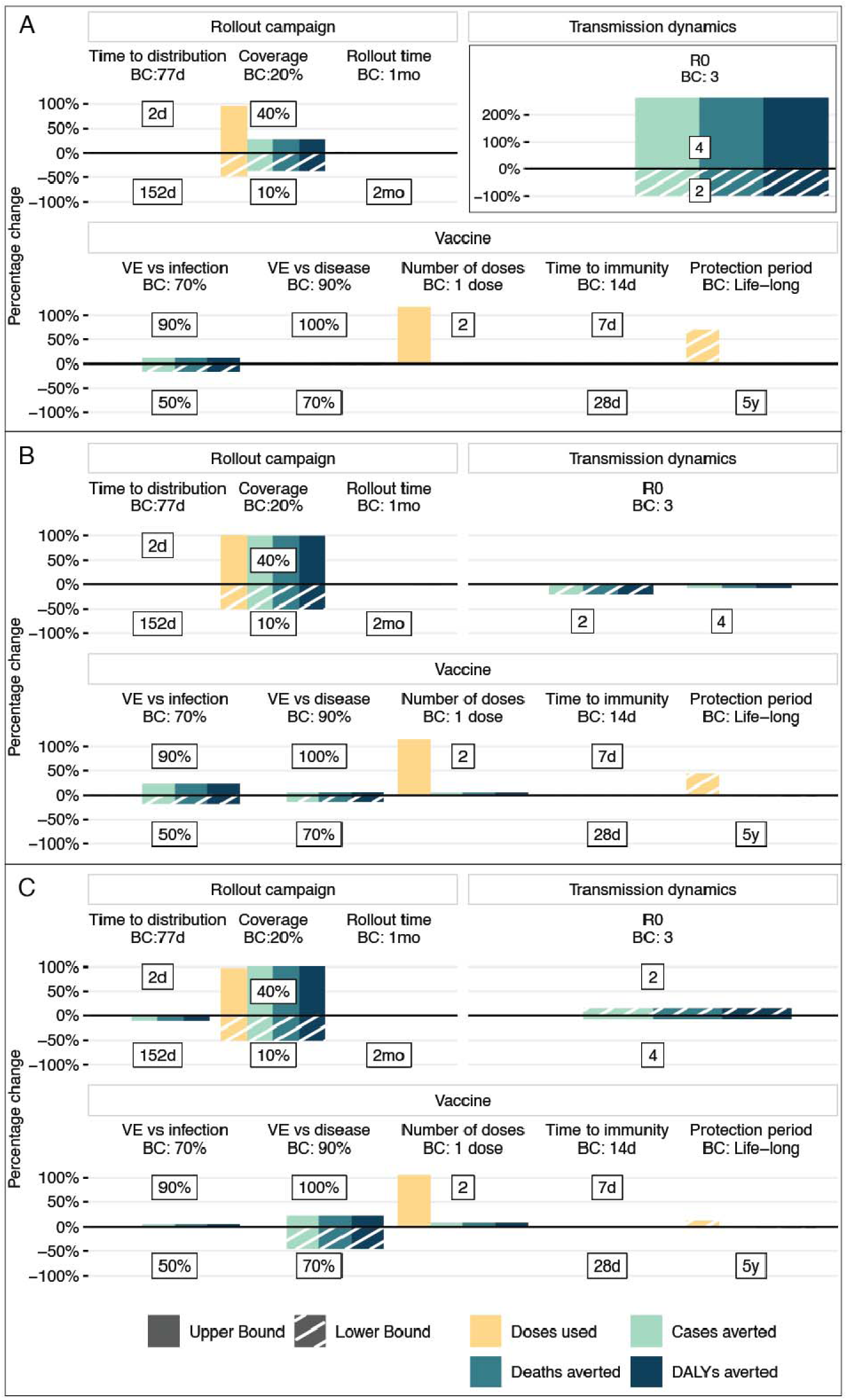
Sensitivity analysis of differences in potential vaccine impact when one parameter changes in comparison to the base case (BC) scenario for reactive vaccination in extended outbreaks with **(A)** high ( ), **(B)** intermediate ( ), and **(C)** no ( ) transmission dampening due to behavioral changes. Comparisons are represented as the percentage change of median estimates of doses used, and cases, deaths, and DALYs averted.

**Figure S5:**
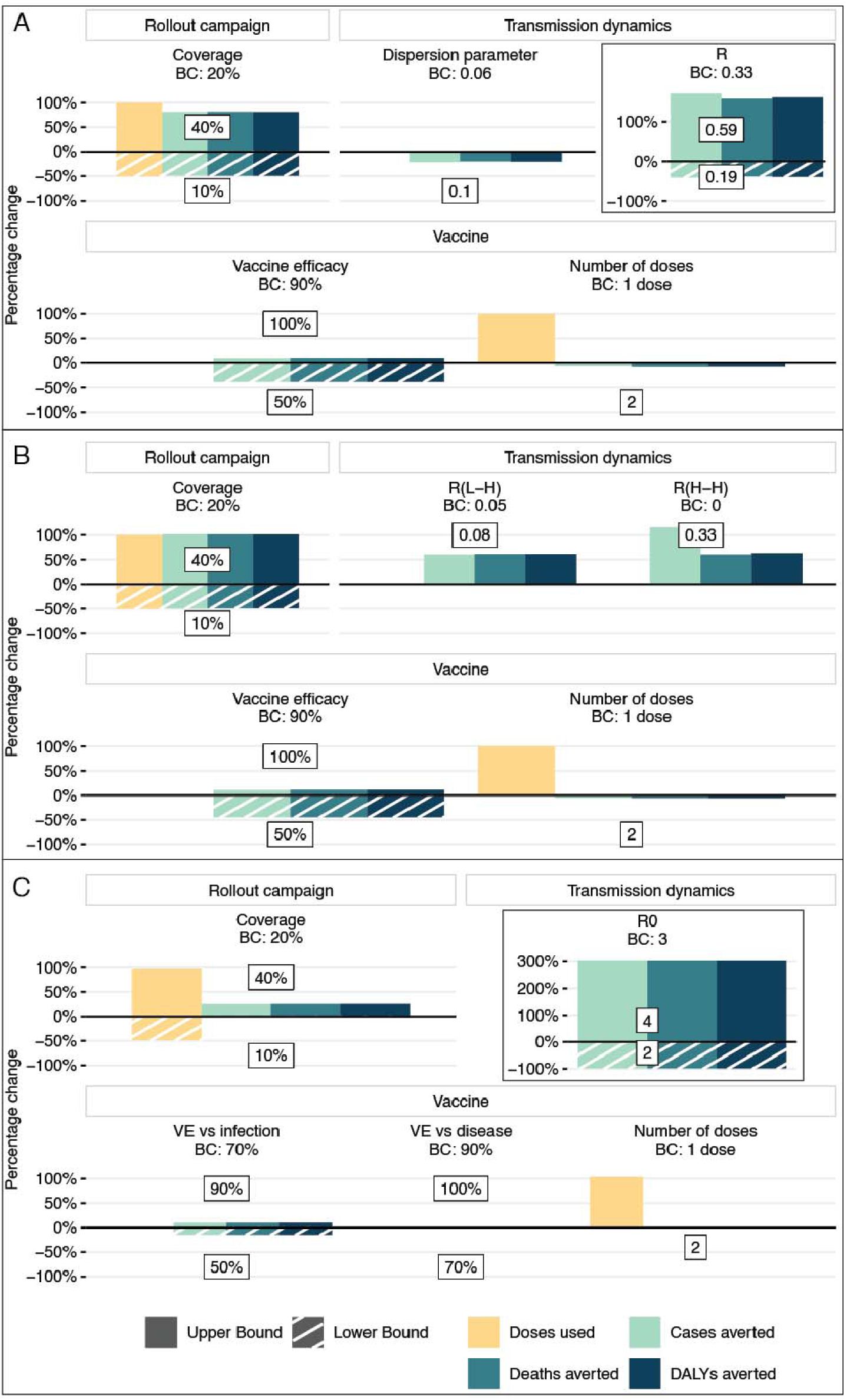
Sensitivity analysis of differences in potential vaccine impact when one parameter changes in comparison to the base case (BC) scenario for proactive vaccination in **(A)** limited, **(B)** livestock amplified, and **(C)** extended outbreaks with a high level of transmission decay due to behavioral changes (*γ* = 0.1). Comparisons are represented as the percentage change of median estimates of doses used, and cases, deaths, and DALYs averted.

**Figure S6:**
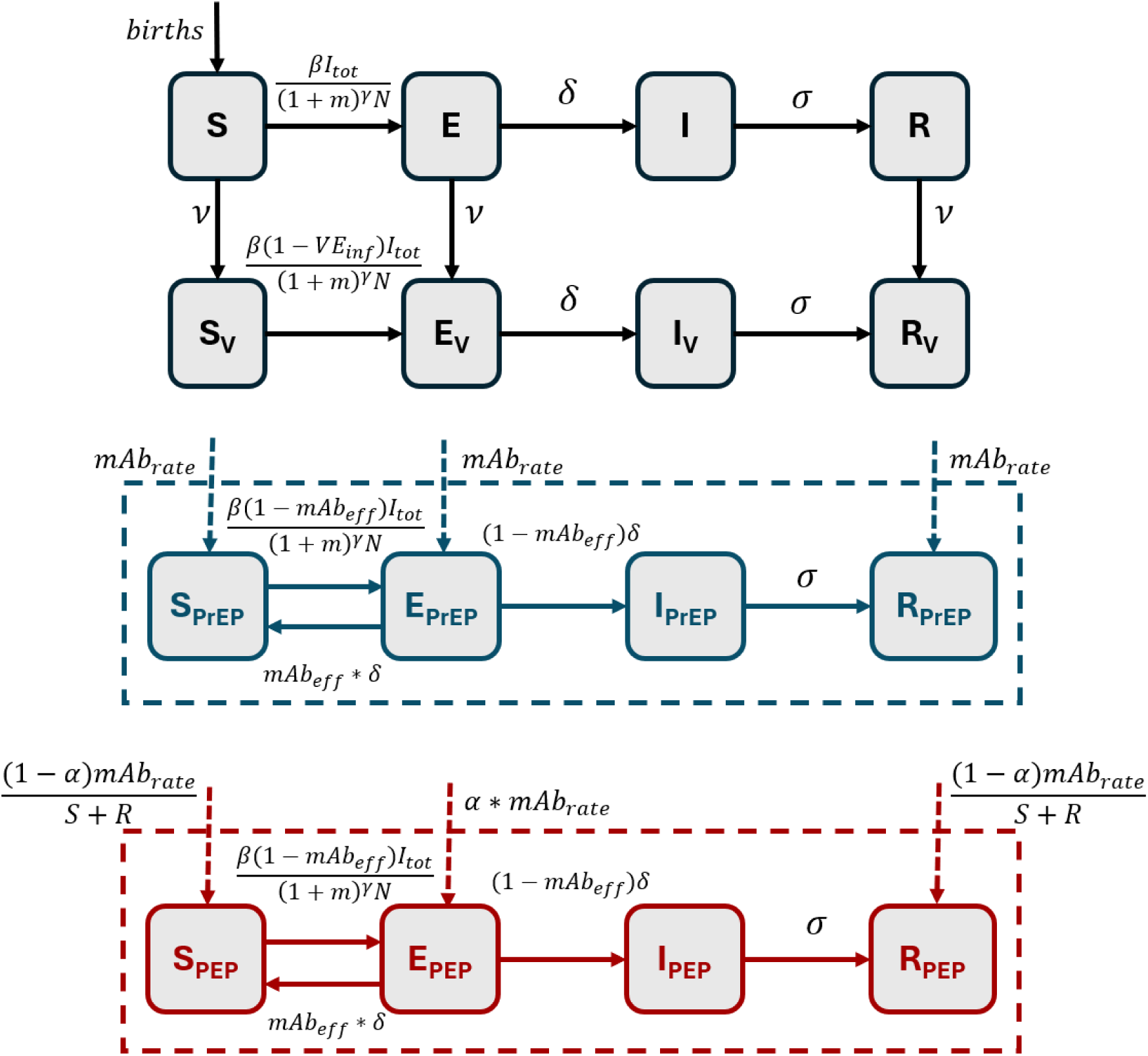
Model structure of stochastic, time-discrete SEIRV model used in extended outbreaks. Natural deaths from all compartments are not represented for simplicity.

**Table S1:**
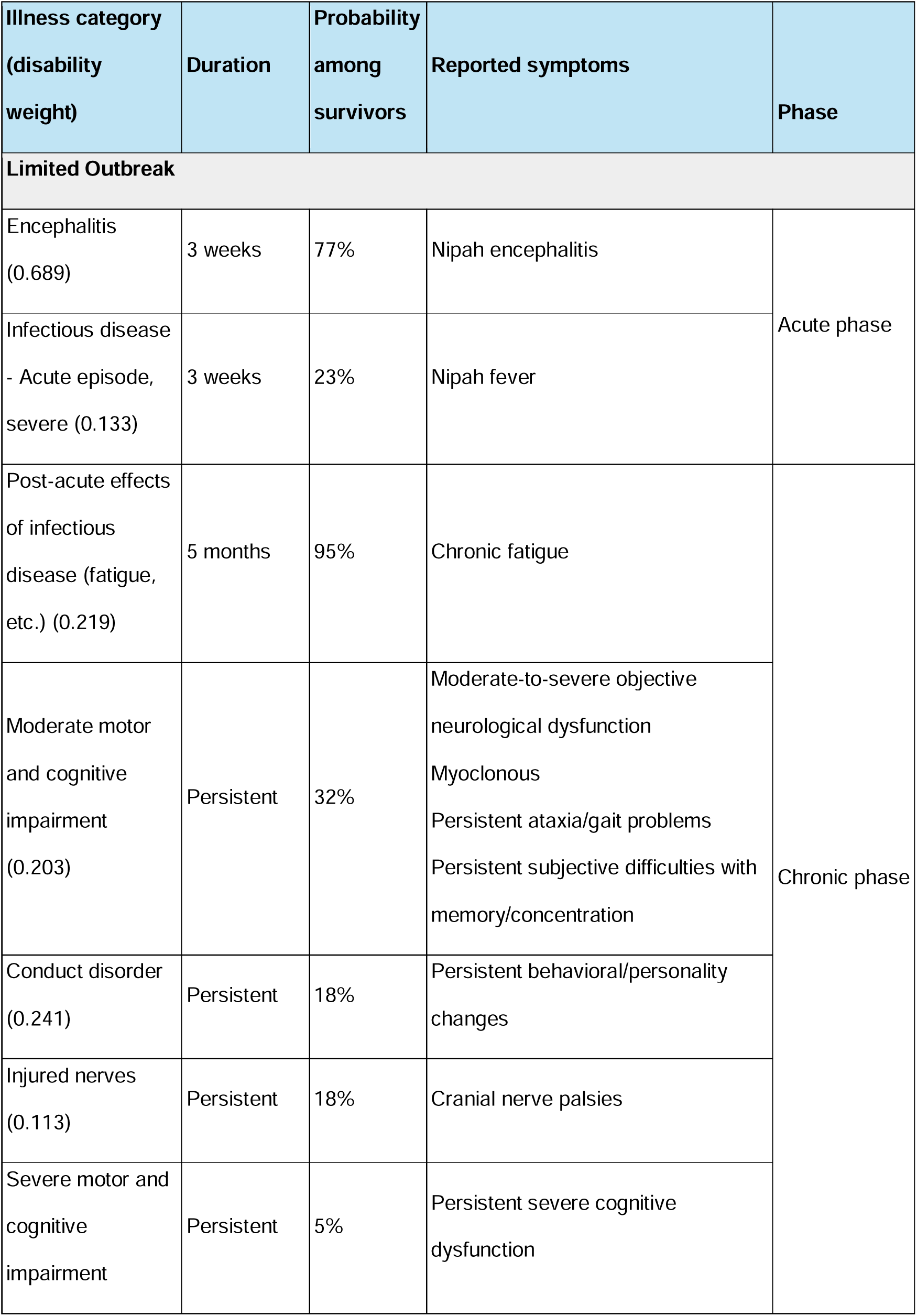
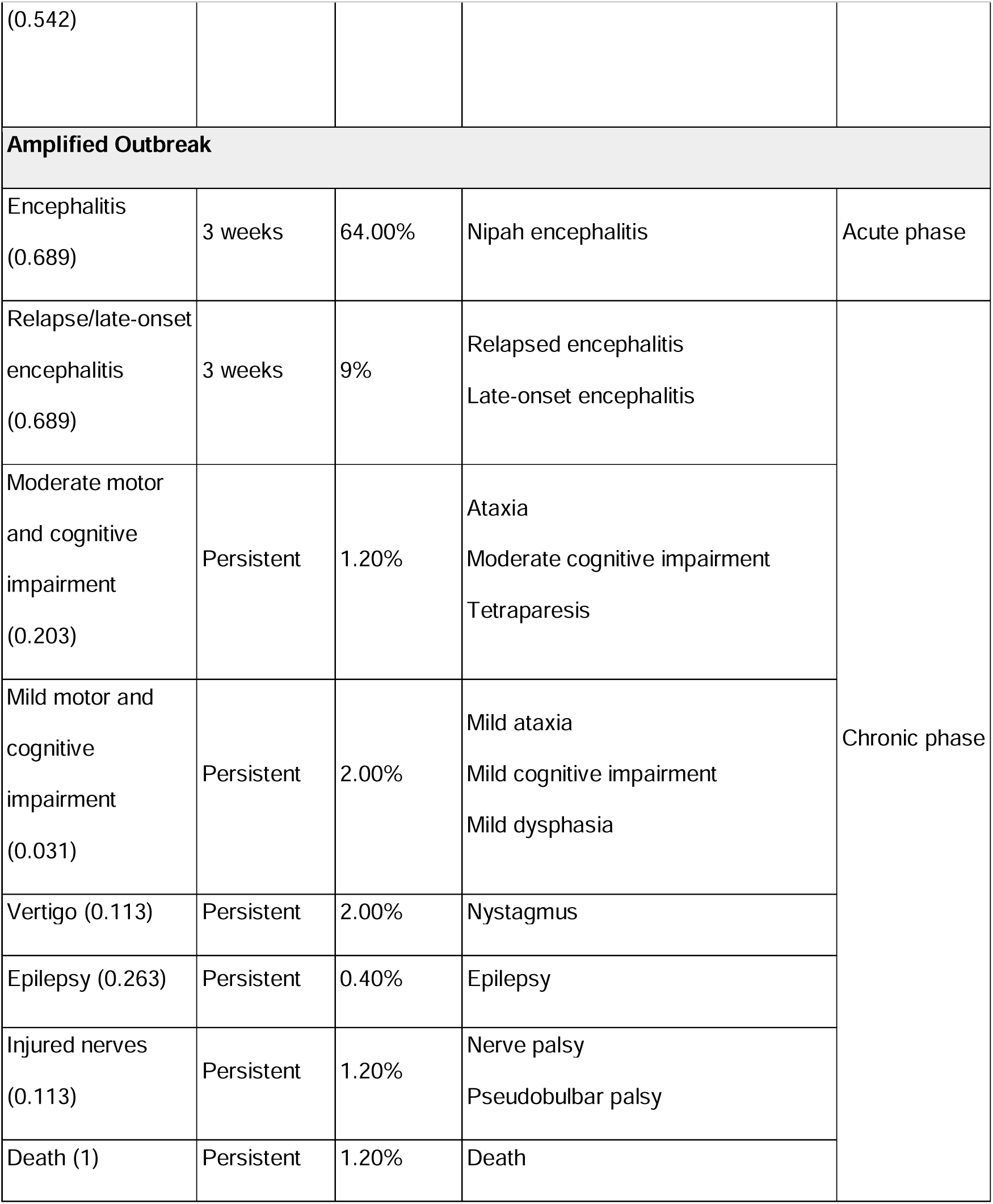
Disability weights, duration and probability among encephalitis survivors of sequelae used for DALY calculations.

## References

1. B. Nikolay, H. Salje, M. J. Hossain, A. K. M. D. Khan, H. M. S. Sazzad, M. Rahman, P. Daszak, U. Ströher, J. R. C. Pulliam, A. M. Kilpatrick, S. T. Nichol, J. D. Klena, S. Sultana, S. Afroj, S. P. Luby, S. Cauchemez, E. S. Gurley, Transmission of Nipah Virus - 14 Years of Investigations in Bangladesh. N. Engl. J. Med. 380, 1804–1814 (2019).

2. P. D. Yadav, K. Baid, D. Y. Patil, T. Shirin, M. Z. Rahman, A. J. Peel, J. H. Epstein, J. M. Montgomery, R. K. Plowright, H. Salje, E. S. Gurley, S. M. Satter, A. Banerjee, A One Health approach to understanding and managing Nipah virus outbreaks. Nat. Microbiol. 10, 1272–1281 (2025).

3. J. H. Epstein, H. E. Field, S. Luby, J. R. C. Pulliam, P. Daszak, Nipah virus: impact, origins, and causes of emergence. Curr. Infect. Dis. Rep. 8, 59–65 (2006).

4. S. M. Satter, W. R. Aquib, S. Sultana, A. R. Sharif, A. Nazneen, M. R. Alam, A. Siddika, F. Akther Ema, K. I. A. Chowdhury, A. N. Alam, M. Rahman, J. D. Klena, M. Z. Rahman, S. Banu, T. Shirin, J. M. Montgomery, Tackling a global epidemic threat: Nipah surveillance in Bangladesh, 2006-2021. PLoS Negl. Trop. Dis. 17, e0011617 (2023).

5. World Health Organization, 2018 Annual Review of Diseases Prioritized under the Research and Development Blueprint. [Preprint] (2018). https://www.who.int/docs/default-source/blue-print/2018-annual-review-of-diseases-prioritized-under-the-research-and-development-blueprint.pdf.

6. S. Kim, H. Kang, L. Skrip, S. Sahastrabuddhe, A. Islam, S.-M. Jung, J. F. Vesga, A. Endo, W. J. Edmunds, K. Abbas, Progress and challenges in Nipah vaccine development and licensure for epidemic preparedness and response. Expert Rev. Vaccines 24, 183–193 (2025).

7. Coalition for Epidemic Preparedness Innovations (CEPI), New vaccine set for human trials in Nipah outbreak hotspot. https://cepi.net//new-vaccine-set-human-trials-nipah-outbreak-hotspot.

8. P. Joi, Nipah vaccines set to enter human trials, GAVI communication (2025). https://www.gavi.org/vaccineswork/nipah-vaccines-enter-human-trials.

9. X. H. S. Chan, I. L. Haeusler, B. J. K. Choy, M. Z. Hassan, J. Takata, T. P. Hurst, L. M. Jones, S. Loganathan, E. Harriss, J. Dunning, J. Tarning, M. W. Carroll, P. W. Horby, P. L. Olliaro, Therapeutics for Nipah virus disease: a systematic review to support prioritisation of drug candidates for clinical trials. Lancet Microbe 6, 101002 (2025).

10. Coalition for Epidemic Preparedness Innovations (CEPI), New human trials for novel antibody offer hope for immediate protection against deadly Nipah. https://cepi.net//new-human-trials-novel-antibody-offer-hope-immediate-protection-against-deadly-nipah.

11. B. Nikolay, G. Ribeiro Dos Santos, M. Lipsitch, M. Rahman, S. P. Luby, H. Salje, E. S. Gurley, S. Cauchemez, Assessing the feasibility of Nipah vaccine efficacy trials based on previous outbreaks in Bangladesh. Vaccine 39, 5600–5606 (2021).

12. B. Nikolay, H. Salje, A. K. M. D. Khan, H. M. S. Sazzad, S. M. Satter, M. Rahman, S. Doan, B. Knust, M. S. Flora, S. P. Luby, S. Cauchemez, E. S. Gurley, A Framework to Monitor Changes in Transmission and Epidemiology of Emerging Pathogens: Lessons From Nipah Virus. J. Infect. Dis. 221, S363–S369 (2020).

13. J. J. Sejvar, J. Hossain, S. K. Saha, E. S. Gurley, S. Banu, J. D. Hamadani, M. A. Faiz, F. M. Siddiqui, Q. D. Mohammad, A. H. Mollah, R. Uddin, R. Alam, R. Rahman, C. T. Tan, W. Bellini, P. Rota, R. F. Breiman, S. P. Luby, Long-term neurological and functional outcome in Nipah virus infection. Ann. Neurol. 62, 235–242 (2007).

14. Centers for Disease Control and Prevention (CDC), Update: Outbreak of Nipah Virus -- Malaysia and Singapore, 1999 (1999). https://www.cdc.gov/mmwr/preview/mmwrhtml/00057012.htm.

15. J. R. C. Pulliam, J. H. Epstein, J. Dushoff, S. A. Rahman, M. Bunning, A. A. Jamaluddin, A. D. Hyatt, H. E. Field, A. P. Dobson, P. Daszak, Henipavirus Ecology Research Group (HERG), Agricultural intensification, priming for persistence and the emergence of Nipah virus: a lethal bat-borne zoonosis. J. R. Soc. Interface 9, 89–101 (2012).

16. S. Cauchemez, S. Epperson, M. Biggerstaff, D. Swerdlow, L. Finelli, N. M. Ferguson, Using routine surveillance data to estimate the epidemic potential of emerging zoonoses: application to the emergence of US swine origin influenza A H3N2v virus. PLoS Med. 10, e1001399 (2013).

17. C. T. Tan, K. J. Goh, K. T. Wong, S. A. Sarji, K. B. Chua, N. K. Chew, P. Murugasu, Y. L. Loh, H. T. Chong, K. S. Tan, T. Thayaparan, S. Kumar, M. R. Jusoh, Relapsed and late-onset Nipah encephalitis. Ann. Neurol. 51, 703–708 (2002).

18. Ministry of Health and Family Welfare of the People’s Republic of Bangladesh, Health Labour Market Analysis in Bangladesh 2021 (2021; https://hsd.portal.gov.bd/sites/default/files/files/hsd.portal.gov.bd/page/e620d076_40f0_4f38_a87a_7897e006a91d/Health%20Labour%20Market%20Analysis%20in%20Bangladesh_2021.pdf).

19. Ministry of Health and Family Welfare-Government of the People’s Republic of Bangladesh, Bangladesh Health Workforce Strategy 2023 - 2041 (2023; https://dghs.portal.gov.bd/sites/default/files/files/dghs.portal.gov.bd/page/b1bc841a_55a4_4d2e_93a9_de32dcb258de/2025-04-09-12-13-9ffe91424690efe99dc6abb2c3653aea.pdf).

20. R. K. Plowright, C. R. Parrish, H. McCallum, P. J. Hudson, A. I. Ko, A. L. Graham, J. O. Lloyd-Smith, Pathways to zoonotic spillover. Nat. Rev. Microbiol. 15, 502–510 (2017).

21. O. Cortes-Azuero, N. Lefrancq, B. Nikolay, C. McKee, J. Cappelle, V. Hul, T. P. Ou, T. Hoem, P. Lemey, M. Z. Rahman, A. Islam, E. S. Gurley, V. Duong, H. Salje, The genetic diversity of Nipah virus across spatial scales. J. Infect. Dis., jiae221 (2024).

22. S. Cauchemez, P. Nouvellet, A. Cori, T. Jombart, T. Garske, H. Clapham, S. Moore, H. L. Mills, H. Salje, C. Collins, I. Rodriquez-Barraquer, S. Riley, S. Truelove, H. Algarni, R. Alhakeem, K. AlHarbi, A. Turkistani, R. J. Aguas, D. A. T. Cummings, M. D. Van Kerkhove, C. A. Donnelly, J. Lessler, C. Fraser, A. Al-Barrak, N. M. Ferguson, Unraveling the drivers of MERS-CoV transmission. Proc. Natl. Acad. Sci. U. S. A. 113, 9081–9086 (2016).

23. R Core Team, R: A Language and Environment for Statistical Computing. R Foundation for Statistical Computing [Preprint] (2024). https://www.R-project.org/.

24. R. FitzJohn, W. Hinsley, odin2: Next generation odin. [Preprint] (2025). https://mrc-ide.github.io/odin2.

25. R. FitzJohn, dust2: Next Generation dust. [Preprint] (2025). https://github.com/mrc-ide/dust2.

26. Food and Agriculture Organization of the United Nations (FAO)-World Health Organization (WHO) Global Forum of Food Safety Regulators, JAPANESE ENCEPHALITIS/ NIPAH OUTBREAK IN MALAYSIA. https://www.fao.org/4/ab455e/ab455e.htm.

27. J. A. Salomon, J. A. Haagsma, A. Davis, C. M. de Noordhout, S. Polinder, A. H. Havelaar, A. Cassini, B. Devleesschauwer, M. Kretzschmar, N. Speybroeck, C. J. L. Murray, T. Vos, Disability weights for the Global Burden of Disease 2013 study. Lancet Glob. Health 3, e712–23 (2015).

28. M. Ock, B. Park, H. Park, I.-H. Oh, S.-J. Yoon, B. Cho, M.-W. Jo, Disability weights measurement for 289 causes of disease considering disease severity in Korea. J. Korean Med. Sci. 34, e60 (2019).

29. L. Zeitlin, R. W. Cross, C. Woolsey, B. R. West, V. Borisevich, K. N. Agans, A. N. Prasad, D. J. Deer, L. Stuart, M. McCavitt-Malvido, D. H. Kim, J. Pettitt, J. E. Crowe Jr, K. J. Whaley, D. Veesler, A. Dimitrov, D. M. Abelson, T. W. Geisbert, C. C. Broder, Therapeutic administration of a cross-reactive mAb targeting the fusion glycoprotein of Nipah virus protects nonhuman primates. Sci. Transl. Med. 16, eadl2055 (2024).

30. H. M. Ong, P. A. S. Ibrahim, C. N. Chong, C. T. Tan, J. P. Schee, M. S. Avumegah, R. G. Román, N. G. Cherian, W. F. Wong, L.-Y. Chang, Malaysia outbreak survivors retain detectable Nipah antibodies and memory B cells after 25 years. J. Infect. 90, 106398 (2025).

